# A qualitative study of Benzodiazepine/Z-drug and Opioid co-use patterns and overdose risk

**DOI:** 10.1101/2024.07.26.24311053

**Authors:** Hannah E. Family, Gabriele Vojt, Hannah Poulter, Chris P. Bailey, Ana Paula Abdala Sheikh, Damiana Cavallo, Sara Karimi, Nick Booth, Peter Da Silva, Louise Aitken, Samantha Stewart, Matthew Hickman, Graeme Henderson, Jenny Scott, Joanna M. Kesten

## Abstract

**Background:** Co-use of benzodiazepines and/or ‘z-drugs’ along with opioids is linked to the rise in drug related deaths (DRD) in the UK. Understanding patterns of co-use could inform harm reduction strategies for reducing DRDs. This study explored how people co-use, including dosages, timings, methods of administration, use of other substances and desired effects sought.

**Methods:** Forty-eight semi-structured interviews across Glasgow in Scotland (n=28), Bristol (n=10) and Teesside (n=10) in England with individuals who co-use illicit and/or prescribed opioids and benzodiazepines/z-drugs were conducted. Eighteen interviews were co-facilitated with qualitatively trained local peer researchers. Interviews were analysed using the Framework method.

**Results:** Six co-use patterns were generated: (1) co-use to aid sleep or come down, (2) curated co-use, opioid agonist therapy (OAT) only (3) morning and evening benzodiazepine doses with opioids throughout the day (4) co-use binges (5) co-use throughout the day, (6) benzodiazepine use throughout the day plus OAT. Patterns one to three reflected more controlled co-use with a focus on self-medicating to give confidence, manage anxiety, promote sleep and come-down from cocaine/ketamine. Patterns four to six involved greater poly-drug use, and less controlled co-use with a focus on seeking euphoria (*“warm glow”, “gouching out”)* or oblivion (to escape untreated mental health conditions and trauma). Patterns two, three, five and six involved daily co-use. People switched between patterns depending on available resources (e.g. finances) or changes to prescriptions (opioids or benzodiazepines). Near-fatal overdoses were reported by participants across all co-use patterns.

Patterns four to six were conceptualised as presenting greater overdose risk due to less controlled co-use and more extensive polydrug use.

**Conclusions:** The patterns identified provide opportunities for future harm reduction strategies, tailoring advice to patterns of use, updated prescribing guidance and policies, and the need for better access to mental health care, for people who co-use benzodiazepines and opioids to reduce DRDs.

## Background

The rise in opioid related deaths in the United Kingdom (UK), especially in Scotland, is a public health crisis(1) – recognised by the creation of the Drug Deaths Taskforce in Scotland and the drug strategy ‘From Harm to Hope’(2) in England. Drug related death (DRD) rates, defined in the UK as, *“a death where the underlying cause is drug abuse or drug dependence or (b) a death where the underlying cause is drug poisoning and where any of the substances controlled under the Misuse of Drugs Act 1971 are involved* (*3*)*.”* The DRD rates in Scotland are the highest in Europe and exceed most countries globally (4). Poly-drug use, particularly of prescribed and illicitly manufactured benzodiazepines, z-drugs (non-benzodiazepine hypnotics e.g. zolpidem, zopiclone) and opioids – is potentially a key driver of the increase in UK DRD (4, 5).

In 2019/20 (the most recent prevalence estimates available) there were 293,863 people in England aged 15 to 64 who used opiates (164,279) or both opiates and cocaine both drugs (129,584), combined, this equates to rate of 8.2 per 1000 general population (6) Treatment data for England shows that 137,965 people were receiving treatment for opioid use in the last year (2023/24), and of these 94% were receiving prescribing interventions. This dataset also showed that 10,447 people in touch with treatment services reported use of benzodiazepines (z-drugs not reported)(7). In Scotland, in 2019/20 47,000 people were estimated to have opioid dependence equating to an estimated prevalence of 13.2 per 1000 general population (8, 9). Of these, 61% were estimated to have received opioid agonist therapy (OAT; including methadone, hydrochloride, buprenorphine, buprenorphine & naloxone and long-acting buprenorphine) at some point in 2019/20 (8, 10).

In England and Wales in 2023 there were 5,448 drug poisoning deaths, 2551 of these involved any type of opioid (except paracetamol compounds): 1,453 (27%) of these involving heroin/morphine, 709 involving methadone (13%), 46 buprenorphine (0,8%) and 512 (9%) of all drug poisonings involved a benzodiazepine and 189 (3%) a z-drug/ zopiclone(11). In 2022 in Scotland there were 1051 DRD; of these 82% involved opioids and 70% of these involved co-use of a benzodiazepine (12). There is an association between the increase in deaths in Scotland and increase in street benzodiazepines (Novel Psychoactive Substance- type benzodiazepines) detected at post-mortem (4). Use of benzodiazepines and z-drugs is common among people who use heroin and opioid agonist treatment (OAT)(13, 14) and poly-drug opioid use has long been the norm in the UK and other countries (15). In some studies co-prescription of opioids and benzodiazepines, or of benzodiazepines for people with an opioid use disorder (OUD) is associated with increased risk of death (16, 17). In the USA and UK primary care benzodiazepine prescription is associated with a two-fold increase in mortality in people with OUD in periods both on and off OAT (16, 17). Research has also pointed to similar trends for z-drugs (18). For people in treatment, the type of OAT prescribed may be important, as Methadone is a full agonist with a long and variable half-life (19) and can potentially cause more prolonged respiratory depression compared to buprenorphine, a partial agonist which on its own has a lower risk of respiratory depression due to its ceiling effect (20). A UK based cohort study showed that hospitalisations for non-fatal overdose were higher amongst people prescribed methadone compared to buprenorphine (20). Whilst theoretical assumptions exist based on the individual pharmacology of each drug and studies such as Domzaridou’s et al (20), it is not currently known, how different types of OAT interact with benzodiazepines and affect respiratory depression, and more broadly, the mechanisms by which any opioid and benzodiazepine and/or z-drug interacts to increase mortality risk.

Research to date has highlighted the complex and varying motives for benzodiazepine or z-drug use among people who use opioids. Benzodiazepines and Z-drugs both work on γ-aminobutyric acid type A receptors (GABA_A_ receptors) causing similar hypnotic and sedative effects but are chemically distinct (21). Co-use of benzodiazepines has been reported to enhance the effects of opioids including heroin and methadone (13, 22–25). People who co-use benzodiazepines and opioids also report further, unintentional opioid use through disinhibition and memory loss caused by benzodiazepines. Whilst others report decreased opioid use through enhancing or prolonging effects of benzodiazepines and reducing symptoms of withdrawal, as well as managing anxiety, depression, and insomnia (22–24, 26). Motivations influence the patterns of benzodiazepine use which may vary from using benzodiazepines prior to, after or at the same time as heroin, methadone and buprenorphine (23, 24, 27). Individuals also express preferences for certain types of benzodiazepines (26). Understanding the interplay between motivation and specific patterns of use in a rapidly changing drug market has the potential to support efforts to reduce drug related harm.

## Methods

This study is part of a larger multidisciplinary research project funded by the Medical Research Council (ref: MR/W029162/1) involving both qualitative and neuropharmacology studies to understand how benzodiazepines and opioids interact to increase the likelihood of fatal overdose. This qualitative interview study aimed to explore the patterns of benzodiazepine and opioid co-use, including preferred combinations, dosages, timings, methods of administration and explore with participants how these factors of co-use affected overdose risk.

### Study design

A qualitative interview study with people who co-used prescribed and/or illicit opioids (e.g. heroin, OAT) and benzodiazepines (e.g. diazepam) or z-drugs (e.g. zopiclone).

### Ethics

Ethical approval for this study was obtained from the Faculty of Health Sciences Committee for Research Ethics, University of Bristol (ref 11906).

### Setting

The study was carried out in three geographical areas: Glasgow (Scotland), Bristol (South-West England); and Teesside (North-East England). Bristol and Teesside were selected as comparison sites to Glasgow which has high rates of opioid and benzodiazepine DRDs. Teesside was selected as a study site as it has high opioid related DRDs but less benzodiazepine and opioid poly-drug use compared to Glasgow, however it does have high rates of z-drug use (28) whereas Glasgow and Bristol do not. Bristol was chosen as it has a high prevalence of opioid use, greater crack cocaine- opioid poly-drug use, but lower than average opioid DRDs.

### Sampling

Participants were recruited from community drug treatment and harm reduction services, women’s recovery groups, homeless outreach residential crisis and stabilisation services. English speaking adults were eligible to participate if they currently - or in the last six months - co-used prescribed and/or illicit opioids (e.g. heroin, OAT) and benzodiazepines (e.g. diazepam) or z-drugs (e.g. zopiclone). Guided by the principle of information power we anticipated a sample size of at least 45 interviews would be sufficient (29).

Following the collaborative and intensive pragmatic qualitative (CLIP-Q) approach (30), we collaborated with peer researchers and staff in local specialist drug services prior to and during the research to facilitate recruitment of people with a range of co-use experiences. Data collection and analysis were concurrent, to monitor and ensure adequacy of the sample. The decision to end data collection was informed by information power principles; the study aim, sample specificity (participant characteristics relating to the phenomenon under study), quality and depth of the data and planned analyses (29).

Peer researchers who were employees or volunteers at recruiting services advertised the study (using posters and information sheets) and worked with staff to identify potential participants. Consent for researchers to contact people interested in participating was obtained. Researchers also regularly visited services to opportunistically recruit.

### Data collection

We conducted face-to-face or telephone interviews at the service people were recruited from or a quiet café. Prior to interviewing, participants re-read the participant information sheet and gave written or verbal informed consent. The socioecological framework (31) formed the conceptual framework for the qualitative work, which includes this analyses, and two further separate analyses exploring motivations for co-use (32) and experiences of harm reduction advice and near fatal overdose experiences (33). The socioecological framework provided a lens to understand when and why people co-use within the context of sociological, cultural, economic and political structures. This conceptual framework guided the development of the interview topic guide (see appendix) explored initiation of co-use, motivations and patterns of co-use, near-fatal overdose experiences and the role of different benzodiazepines or Z-drugs and opioids in the person’s life, how the risks of co-use were managed and the characteristics of valuable interventions. The interviews also captured socio-demographics, drug treatment history, health conditions and other relevant social influences in relation to the risks of co-use. Images of illicit benzodiazepines, z-drugs and opioids submitted to WEDINOS (see https://www.wedinos.org/) for drug testing and locally generated testing data were also used as prompts.

Interviews were conducted by HF, HP or GV. Eighteen of these were co-facilitated with a peer researcher. Participants were anticipated to feel more comfortable in the presence of a peer researcher, and they brought valuable insights, explained subtle contexts and probed for details that might otherwise be missed by the university researcher due to their lived experience (34, 35). The number of co-facilitated interviews was determined by the availability of the peer researcher(s), the consent of the participant and host organisation. Peer researchers received interviewing training adapted from (36) and project specific information (e.g. topic guide). The peer researcher asked the main questions, developing a rapport with the participant, and the university researcher asked for further detail as appropriate. Participants received £10 cash or vouchers, depending on the policy of the recruiting organisation, as a token of thanks. The university researcher debriefed participants and peer researchers separately. Interviews were recorded, transcribed (intelligent verbatim) and anonymised.

### Reflexivity

This project was conducted by a multidisciplinary team of researchers with additional clinical (JS) voluntary sector support roles (GH & HP) and expertise in opioid pharmacology (GH, CB, APAS, DC, SK). HF, GV, HP who have extensive qualitative and behavioural science expertise conducted the interviews and led the analysis with support from JS, GH and JK. Following the CLIP-Q approach; a written summary and reflexive notes were made after each interview to inform future interviews, and weekly team meetings held to facilitate a continuous process of reflection, translation and triangulation between each study component. The credibility, integrity and applicability of findings were checked with peer researchers (with lived experience of co-use) and expert stakeholders in each study location at the analysis stage.

### Analysis

‘Framework’ (37) was the analytical method chosen to identify patterns of co-use, it was particularly useful for exploring how individual participant’s co-use may be related to near fatal overdose experiences as well as patterns across the whole dataset. Framework is a qualitative analysis approach that involves the generation of themes, but unlike other thematic approaches to analysis (e.g. reflexive thematic analysis, grounded theory) it includes an extra step where data is charted, between coding (or indexing the data) and the analysis (creating themes and sub-themes). What this adds to other thematic approaches, is the ability to summarise at the individual and group level. According to Ritchie and Spencer (37) reviewing of the charts can also generate analytical insights, in the form of defining concepts, mapping the range and nature of phenomena, creating typologies, finding associations, providing explanations or describing strategies.

For this study, we carried out the Framework analysis as follows. First we (HF, GV and HP) read and re-read transcripts and discussing with the wider team the patterns identified, and coding participant’s descriptions of co-use and near-fatal overdose experiences using NVivo (Release 1.7.2 (1560)). Using a matrix to chart the information for each participant, details of current and past co-use of prescribed and illicit drugs were identified, with as much detail on chronological order of administration, dose and/or quantity as reported, overdose experiences, whether this overdose occurred using their current pattern of co-use and if it had involved naloxone or any medical intervention. The charts were then reviewed, discussed with the wider team, and further details added until it was felt that they captured all the relevant data on how people co-used. The next step involved identifying what themes – or “co-use patterns” were present across the matrix, naming these patterns, and agreeing which participants’ accounts related to the patterns identified. As the focus of this analysis is on patterns of behaviour, the analysis presented in this paper, is intentionally ‘near the data’ and descriptive (38, 39) to provide a detailed picture of how people co-use, in order to inform policy and practice related to co-prescribing and wider harm reduction efforts to address DRD among people who co-use. In addition, the data is informing the neuropharmacology experiments running alongside the interviews, that were testing the mechanisms underlying fatal overdoses. We have also been intentionally descriptive in the naming of the patterns in order that they highlight the specificity of which types of drugs are co-used and when during the day. Further papers (33, 40) present more interpretive analyses of the qualitative data, and capture the motivations for, and experiences of co-use, as well as the socioecological context that co-use occurs in, and the support available to them, and the harm reduction strategies they have available to keep themselves safe.

Guided by recommendations on quantification of qualitative data (41), the numbers of participants whose current co-use mapped to each pattern and overdose experience are reported. We have mapped participants to their most current and frequently used pattern. No inferences can or should be drawn about the prevalence of the patterns amongst people who co-use beyond the sample in this study.

## Results

Between November 2022-September 2023, 47 face-to-face and 1 telephone interview were conducted, lasting between 20-103 (mean 49.6) minutes. Participant demographics for the whole sample are reported in Table 1, and according to pattern in Table 4. The participants predominantly self-identified as White British / Scottish or English, 35 (73%) of our participants self-identified as male, and our participants ranged in age from 25-61 years of age. Twenty-eight interviews were conducted with people from Glasgow, 10 from Teesside, and 10 from Bristol. Forty-three (90%) of our participants were prescribed OAT or other opioids (methadone n=27, buprenorphine n=9, long-acting injectable buprenorphine n=6, morphine n=1, dihydrocodeine n=1, heroin assisted treatment n=1).

**Table 1.**
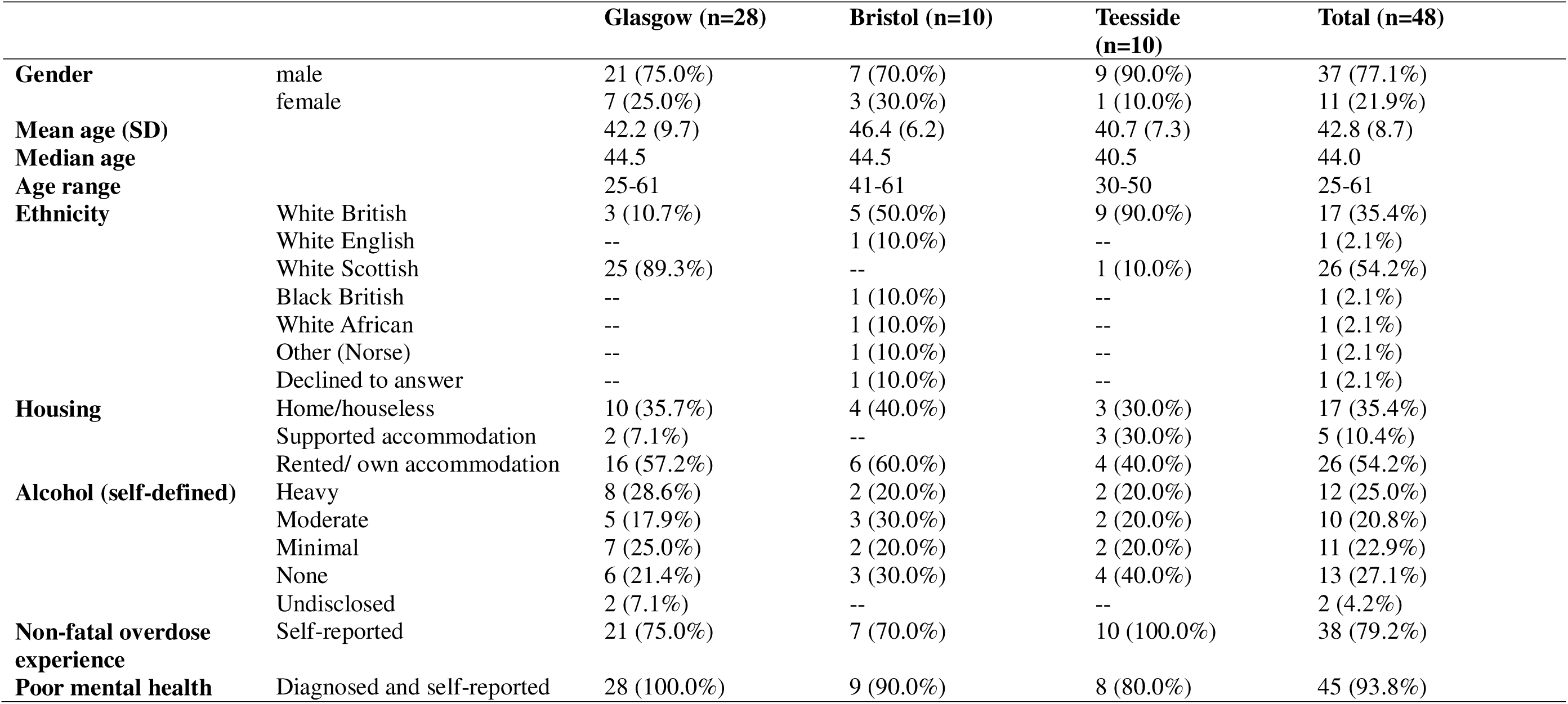
Participant demographics for the total sample and per study location

|  |  | Glasgow (n=28) | Bristol (n=10) | Teesside (n=10) | Total (n=48) |
| --- | --- | --- | --- | --- | --- |
| **Gender** | male | 21 (75.0%) | 7 (70.0%) | 9 (90.0%) | 37 (77.1%) |
|  | female | 7 (25.0%) | 3 (30.0%) | 1 (10.0%) | 11 (21.9%) |
| **Mean age (SD)** |  | 42.2 (9.7) | 46.4 (6.2) | 40.7 (7.3) | 42.8 (8.7) |
| **Median age** |  | 44.5 | 44.5 | 40.5 | 44.0 |
| **Age range** |  | 25-61 | 41-61 | 30-50 | 25-61 |
| **Ethnicity** | White British | 3 (10.7%) | 5 (50.0%) | 9 (90.0%) | 17 (35.4%) |
|  | White English | -- | 1 (10.0%) | -- | 1 (2.1%) |
|  | White Scottish | 25 (89.3%) | -- | 1 (10.0%) | 26 (54.2%) |
|  | Black British | -- | 1 (10.0%) | -- | 1 (2.1%) |
|  | White African | -- | 1 (10.0%) | -- | 1 (2.1%) |
|  | Other (Norse) | -- | 1 (10.0%) | -- | 1 (2.1%) |
|  | Declined to answer | -- | 1 (10.0%) | -- | 1 (2.1%) |
| **Housing** | Home/houseless | 10 (35.7%) | 4 (40.0%) | 3 (30.0%) | 17 (35.4%) |
|  | Supported accommodation | 2 (7.1%) | -- | 3 (30.0%) | 5 (10.4%) |
|  | Rented/ own accommodation | 16 (57.2%) | 6 (60.0%) | 4 (40.0%) | 26 (54.2%) |
| **Alcohol (self-defined)** | Heavy | 8 (28.6%) | 2 (20.0%) | 2 (20.0%) | 12 (25.0%) |
|  | Moderate | 5 (17.9%) | 3 (30.0%) | 2 (20.0%) | 10 (20.8%) |
|  | Minimal | 7 (25.0%) | 2 (20.0%) | 2 (20.0%) | 11 (22.9%) |
|  | None | 6 (21.4%) | 3 (30.0%) | 4 (40.0%) | 13 (27.1%) |
|  | Undisclosed | 2 (7.1%) | -- | -- | 2 (4.2%) |
| **Non-fatal overdose experience** | Self-reported | 21 (75.0%) | 7 (70.0%) | 10 (100.0%) | 38 (79.2%) |
| **Poor mental health** | Diagnosed and self-reported | 28 (100.0%) | 9 (90.0%) | 8 (80.0%) | 45 (93.8%) |

Twenty participants (42%) were currently prescribed benzodiazepines (diazepam n=19, nitrazepam n =1) (see Table 2).

**Table 2:**
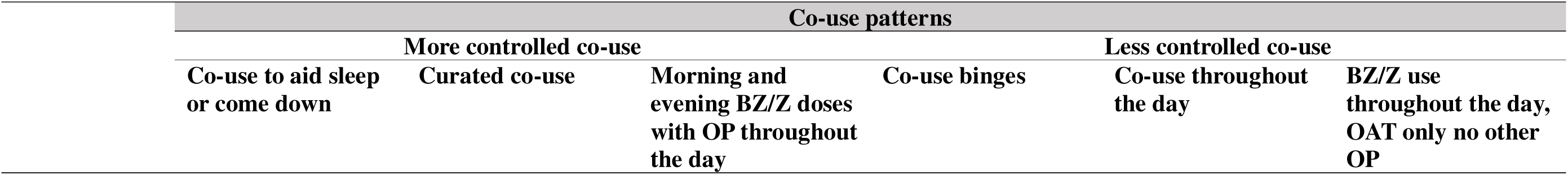
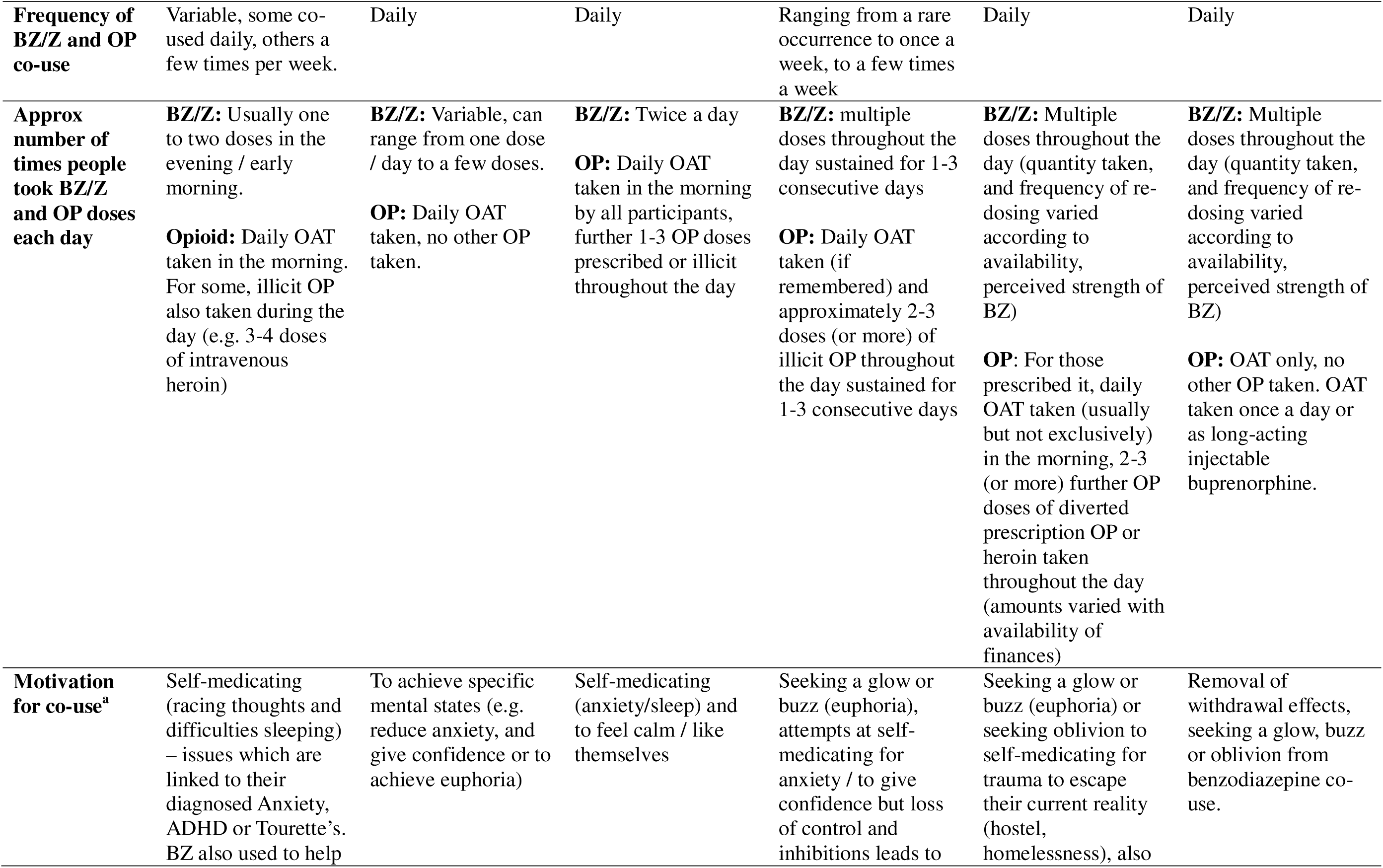
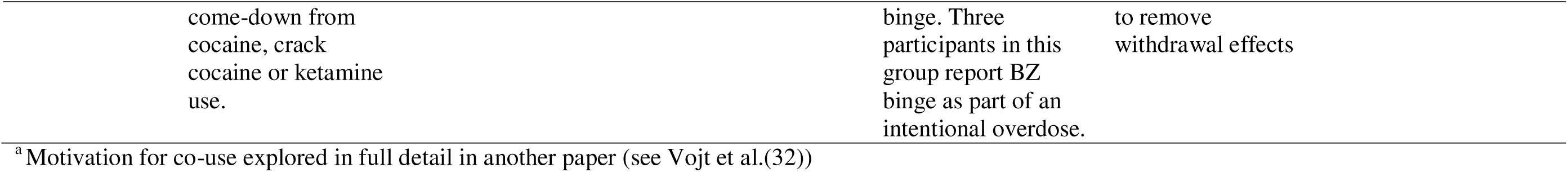
Summary of co-use patterns, motivations for co-use and approximate numbers of benzodiazepine (BZ), z-drug (Z) and opioid (OP) doses per day

Participant’s descriptions of co-use included rich details around their reasons for co-use, the purpose it served in their lives, factors that empowered or constrained their own attempts to co-use safely or reduce their co-use and how their patterns of co-use varied across their life-course and with changes in illicit and licit drug markets.

All participants described how their current co-use patterns were influenced by intrapersonal factors (e.g. mental health symptoms, availability of finances, unstable housing), interpersonal factors (family dynamics, relationships, loss and trauma) and organisational and system level factors (e.g. availability of licit and illicit opioids and benzodiazepines). Finances, housing, access to prescriptions and illicit drug markets were variable, and for this reason, participants often described following more than one pattern.

### 1. Six patterns of co-use

Six patterns of co-use were identified (Table 2). The first three patterns exemplify more controlled co-use, through restricted use of benzodiazepines and opioids, with doses of benzodiazepines timed to achieve a specific function (e.g. sleep, manage mental health symptoms): (1) co-use to aid sleep or come down, (2) curated co-use, OAT only, no other opioids (3) morning and evening benzodiazepine doses with opioids throughout the day. In these first three patterns, doses of benzodiazepines and opioids were taken separately from each other at different times of day. However, it is classed as co-use due to overlapping lengths of time each drug is active in the body. The other three patterns, involve less controlled co-use with higher doses and greater polydrug use, and 70% (n=34) of our participants reported currently following one of these patterns, which were: (4) co-use binges, (5) co-use throughout the day, (6) benzodiazepines/z-drugs throughout the day plus OAT (no other opioid), where use of benzodiazepines and opioids can be simultaneous / timed very closely (e.g. ranging from 5-60 mins between doses). Table 3 describes the approximate daily quantities of prescribed and illicit benzodiazepines and opioids and other prescribed and illicit drugs taken. Table 4 reports the numbers of participants currently using each pattern and sociodemographic data. Nineteen percent (n=9 (4 (45%) were male)) of our participants reported currently following the more controlled co-use patterns (patterns 1-3) and 70% (n=34 (26 (77%) were male)) reported currently following one of the less controlled co-use patterns (patterns 4-6) (Table 4). Age ranges were similar for all patterns.

**Table 3:**
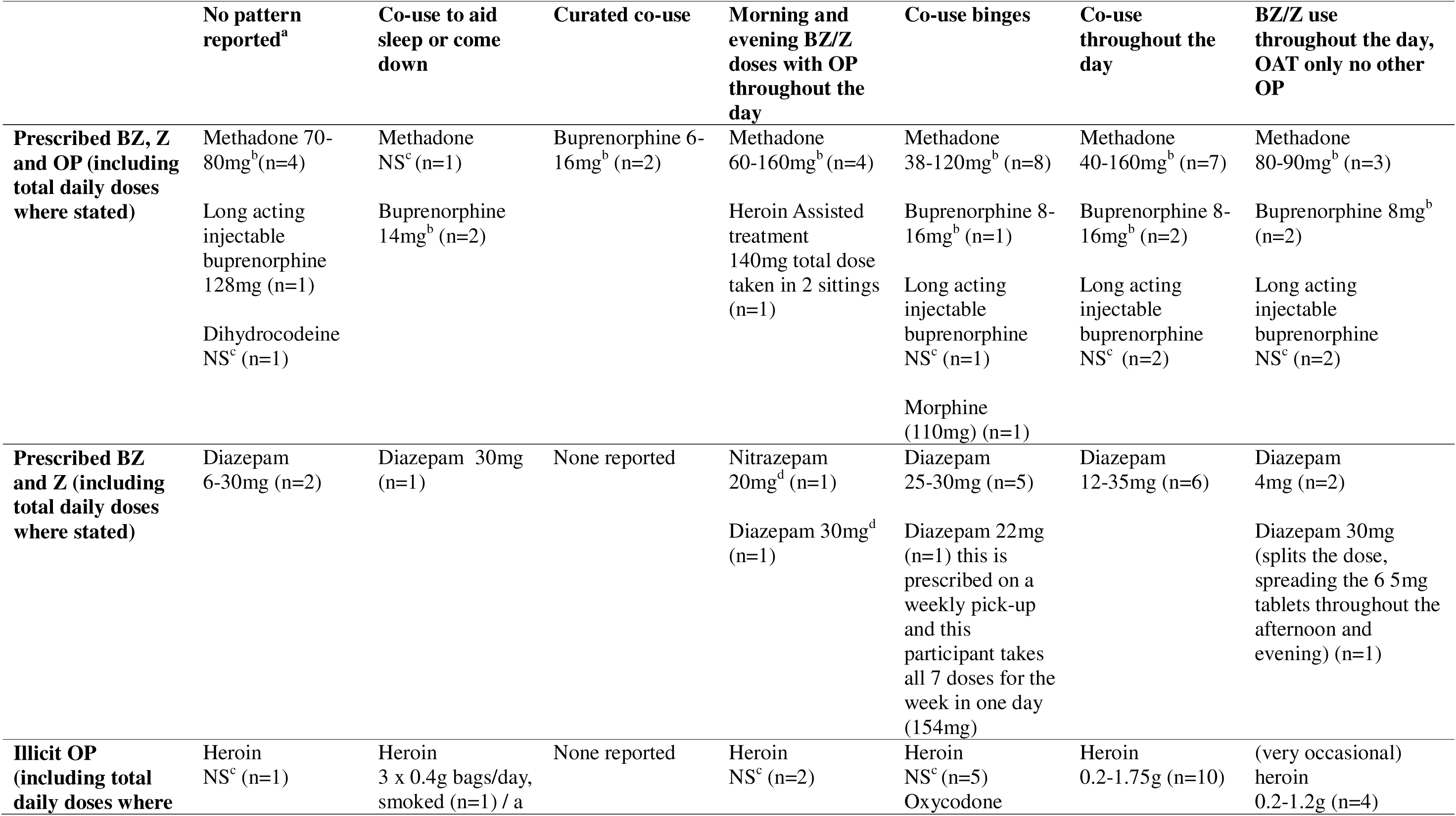
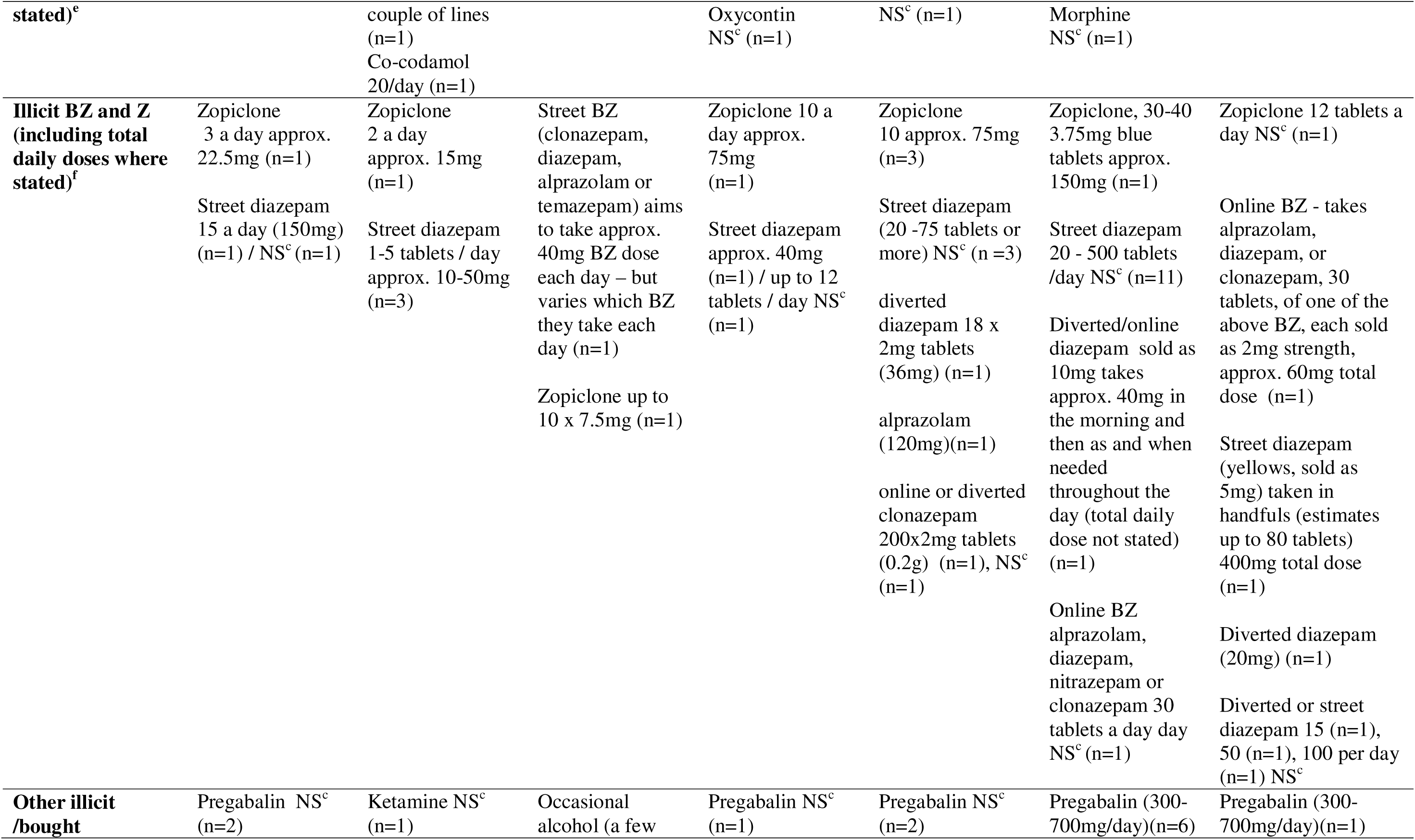
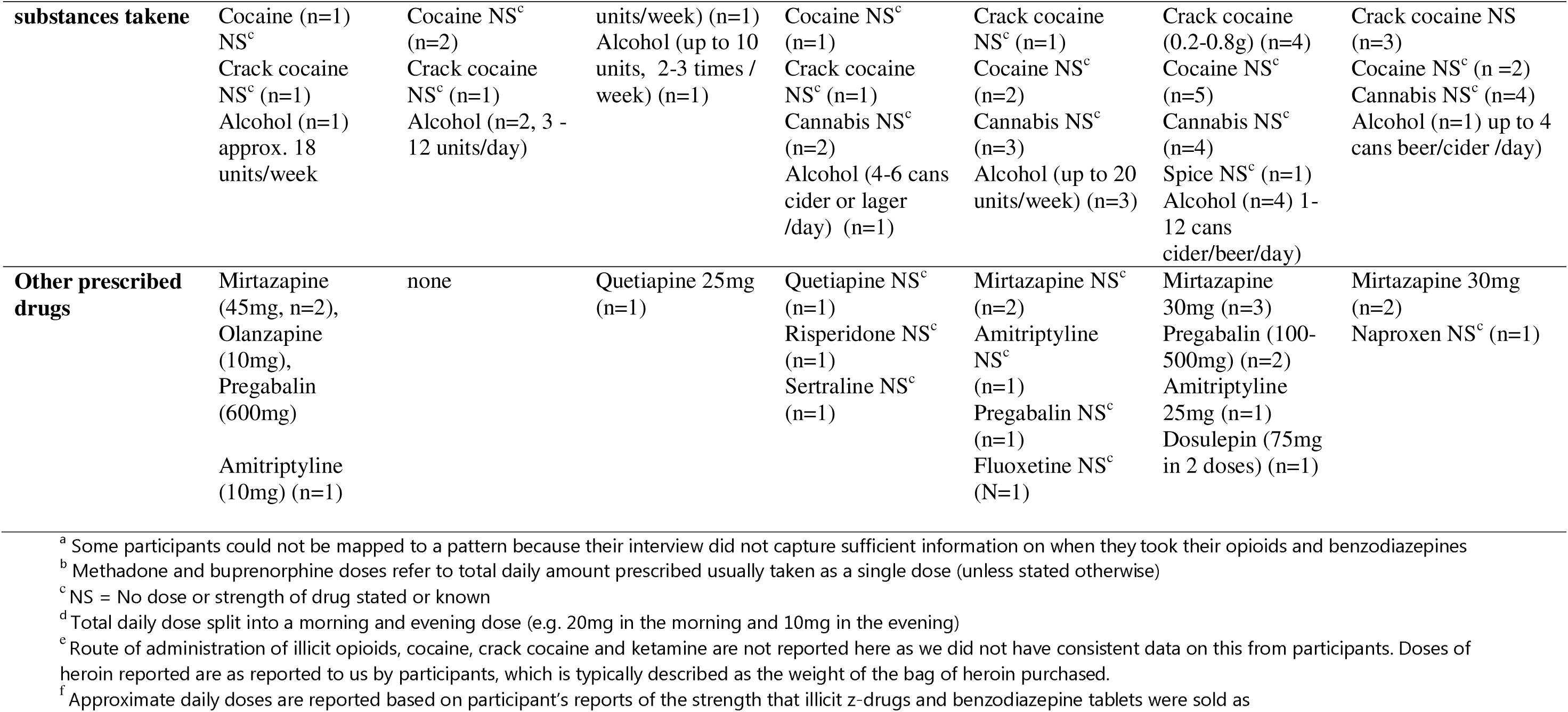
Approximate quantities of prescribed and illicit benzodiazepine and opioid used, other prescribed and illicit medication taken reported by participants in each co-use pattern

**Table 4:**
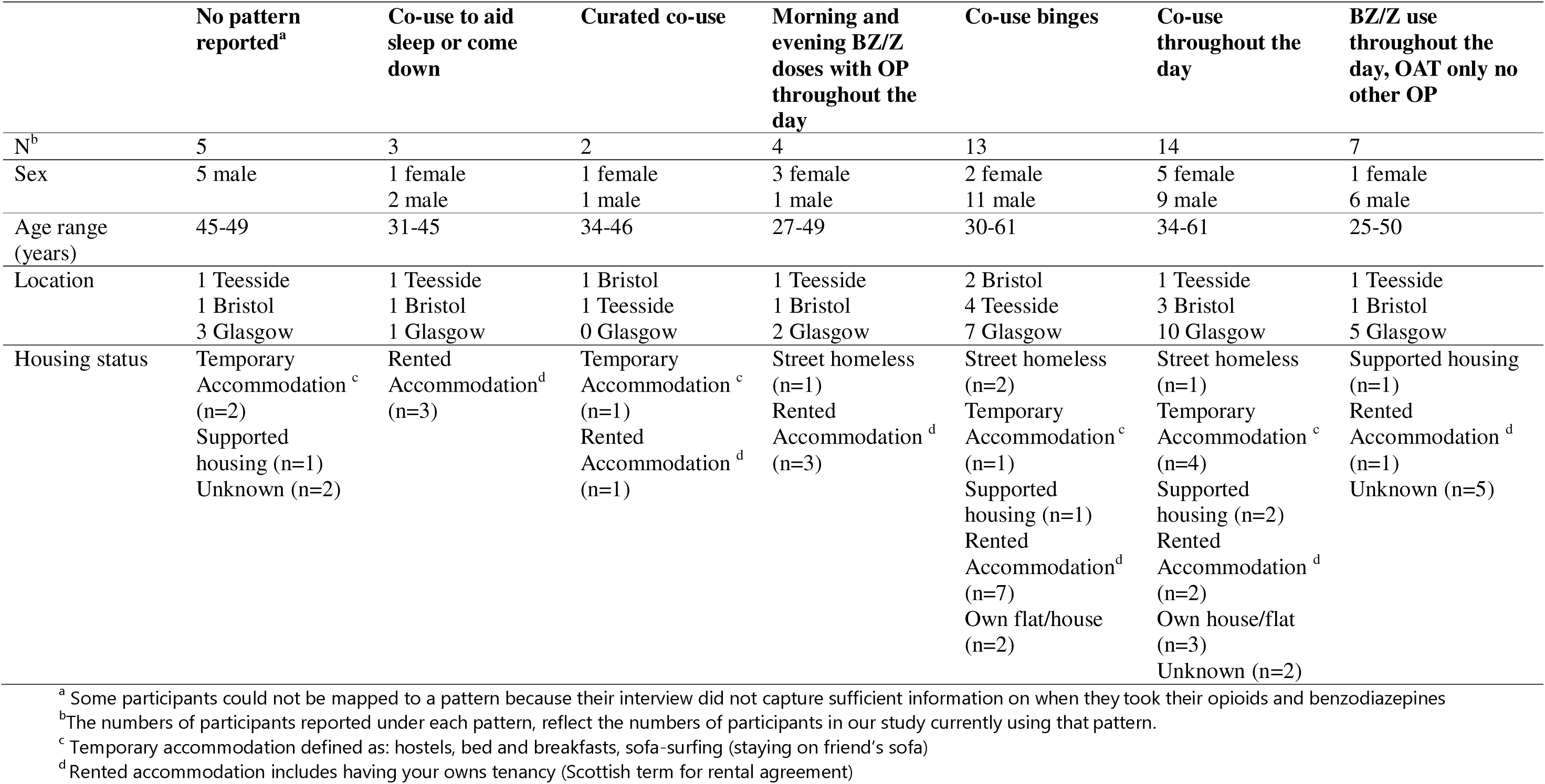
Co-use patterns, numbers of people who reported currently using this co-use pattern and participant demographics

|  | **No pattern reported<sup>a</sup>** | **Co-use to aid sleep or come down** | **Curated co-use** | **Morning and evening BZ/Z doses with OP throughout the day** | **Co-use binges** | **Co-use throughout the day** | **BZ/Z use throughout the day, OAT only no other OP** |
| --- | --- | --- | --- | --- | --- | --- | --- |
| N<sup>b</sup> | 5 | 3 | 2 | 4 | 13 | 14 | 7 |
| Sex | 5 male | 1 female<br>2 male | 1 female<br>1 male | 3 female<br>1 male | 2 female<br>11 male | 5 female<br>9 male | 1 female<br>6 male |
| Age range (years) | 45-49 | 31-45 | 34-46 | 27-49 | 30-61 | 34-61 | 25-50 |
| Location | 1 Teesside<br>1 Bristol<br>3 Glasgow | 1 Teesside<br>1 Bristol<br>1 Glasgow | 1 Bristol<br>1 Teesside<br>0 Glasgow | 1 Teesside<br>1 Bristol<br>2 Glasgow | 2 Bristol<br>4 Teesside<br>7 Glasgow | 1 Teesside<br>3 Bristol<br>10 Glasgow | 1 Teesside<br>1 Bristol<br>5 Glasgow |
| Housing status | Temporary Accommodation <sup>c</sup> (n=2)<br>Supported housing (n=1)<br>Unknown (n=2) | Rented Accommodation<sup>d</sup> (n=3) | Temporary Accommodation <sup>c</sup> (n=1)<br>Rented Accommodation <sup>d</sup> (n=1) | Street homeless (n=1)<br>Rented Accommodation <sup>d</sup> (n=3) | Street homeless (n=2)<br>Temporary Accommodation <sup>c</sup> (n=1)<br>Supported housing (n=1)<br>Rented Accommodation<sup>d</sup> (n=7)<br>Own flat/house (n=2) | Street homeless (n=1)<br>Temporary Accommodation <sup>c</sup> (n=4)<br>Supported housing (n=2)<br>Rented Accommodation <sup>d</sup> (n=2)<br>Own house/flat (n=3)<br>Unknown (n=2) | Supported housing (n=1)<br>Rented Accommodation <sup>d</sup> (n=1)<br>Unknown (n=5) |

<sup>a</sup> Some participants could not be mapped to a pattern because their interview did not capture sufficient information on when they took their opioids and benzodiazepines
<sup>b</sup> The numbers of participants reported under each pattern, reflect the numbers of participants in our study currently using that pattern.
<sup>c</sup> Temporary accommodation defined as: hostels, bed and breakfasts, sofa-surfing (staying on friend's sofa)
<sup>d</sup> Rented accommodation includes having your own tenancy (Scottish term for rental agreement)

### 1 [Insert **tables 2, 3 and 4** here]Co-use to aid sleep or come down (n=3)

Prescribed OAT was taken in the morning, with variable amounts of illicit opioids throughout the day. A key feature, not reported in patterns 2 and 3, was regular use of stimulants (cocaine, crack cocaine, ketamine). Benzodiazepines or z-drugs were only taken at night time, taken to promote sleep; to help calm racing thoughts (more frequent benzodiazepine use), or to come down from illicit stimulant (ketamine or cocaine) taken on a night out. Unlike patterns 2 and 3, co-use did not happen in the daytime, and co-use was described as more occasional, and only for functional reasons - after a night out to come down from stimulants taken, or for nights when their thoughts were racing (managing mental health symptoms) so that they could sleep. Figure 1 and the quote below gives an example co-use timeline for this pattern, reported by participant 31 (female, Glasgow).

> *“First thing in the morning, as soon I get up in the morning, I take the medication* [8mg buprenorphine] *under my tongue…*[later she said] *I was taking them* [diazepam] *for a cocaine* [powdered, snorted] *comedown, rather than taking them as a medication. So, when I was trying to get to sleep, I would use them* [diazepam] *to come off.”* (P31, Female, Glasgow)

**Figure 1:**
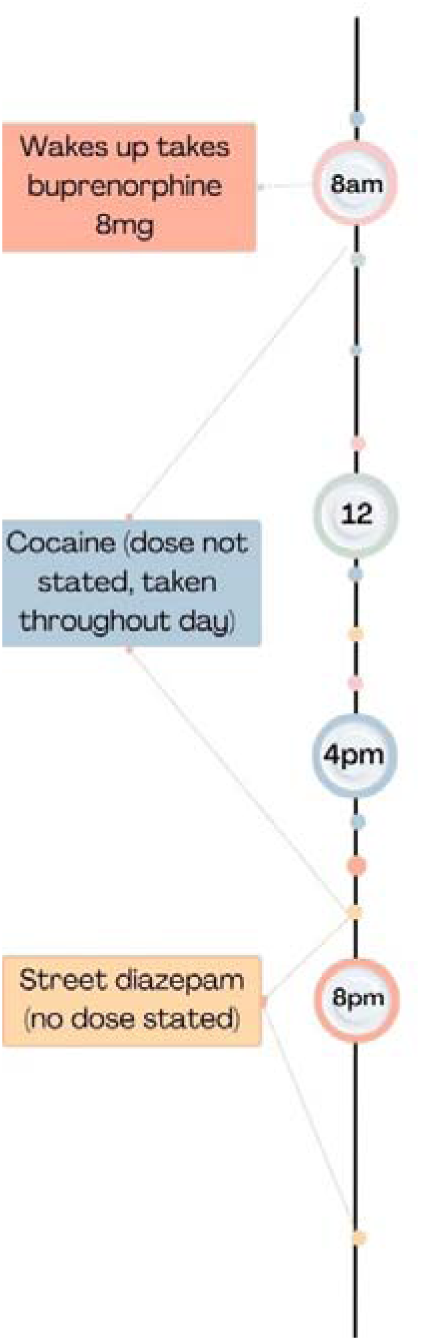
Timeline of benzodiazepine and opioid co-use from participant 31 who reported co-use as a tool to come down from cocaine

All participants currently following this pattern reported diagnosed but untreated mental health conditions and using benzodiazepines to stop their *“mind racing”* (P25, Male, Teesside). One participant had a prescription for 30mg diazepam. Others had had requests for prescribed benzodiazepines to help with their anxiety and sleep declined, so instead used illicit benzodiazepines (approximately 1-5 tablets, 10-50mg) sourced on the street or online. Participants described exerting restraint over their benzodiazepine use, limiting it to evening use and to the minimum effective dose (for them) to reduce the risk of unwanted behaviour and developing tolerance to high doses:

> *“As little as possible. Just 10mg to get me to sleep at night* […] *sometimes I ended up getting a bit too involved* [with benzodiazepines]*– building up tolerance and taking too many – and then I start getting into trouble.”* (P14, Male, Bristol)

Participant 25 (Male, Teesside) reported a near-fatal overdose through this co-use pattern that resulted in him being hospitalised and in a coma for over a month. He explained that it was caused by the variable strength of a batch of MSJs [one type of illicit diazepam] he took one evening.

> “*I’d been getting that particular batch of MSJs for about three days and the normal dose I was taking two on a dinnertime and two on a night, they weren’t doing the job* […] *So then I would [normally] just like increase by five* […] *but on this particular time I took 15 in one go* […] *you don’t know what’s in them, you really don’t. So it’s hit and miss sometimes, you could get a batch that’s really strong, sometimes you get a batch that’s really weak.* (P25, Male, Teesside).

Through provision of visual images alongside local testing data during the interview we were able to ascertain that ‘MSJ’ tablets circulating in Teesside contained bromazolam, which has been linked to drug related deaths throughout England and Wales (42).

### 2 Curated co-use (n=2)

Another pattern involved carefully curating co-use each day varying timing, doses, combinations or choosing different benzodiazepines or z-drugs to achieve specific mood states. In this pattern, co-use was intentional, like pattern one, to serve a function – including reducing social anxiety, increasing confidence to present at work (lower doses of benzodiazepines / z-drugs taken throughout the day). Unlike pattern 1, also for experiential reasons – to achieve euphoria (described as a warm glow, or buzz) (involving higher doses of benzodiazepines/z-drugs taken at once). Both participants who reported this pattern were prescribed buprenorphine but did not have a benzodiazepine/z-drug prescription and so sourced online, street or diverted versions. Unlike patterns 1 and 3, in this pattern, illicit opioids were not taken on-top of their buprenorphine dose and participants did not report any other polydrug use. Participant 5 describes below, and depicted in figure 2, how he curated his co-use:

> *“The reason for why I want to take them* [zopiclone] *influences the time of me taking them. The example I use again will be yesterday’s. I bought them on the evening, and I took them on the evening because there was an event going on, on the phone game I play, where I knew there was going to be a few people talking so I took a few to be social on the evening then*.

> *Whereas if… I’m going to say, that’s on the other hand to like maybe tomorrow or the next day, I don’t know. I might have a few appointments going on in the day. I might feel a little bit shitty so I might go pick some up during the day and then I’d have two before the appointment and then two after the appointment. And then repeat.”* (P5, Male, Teesside).

**Figure 2:**
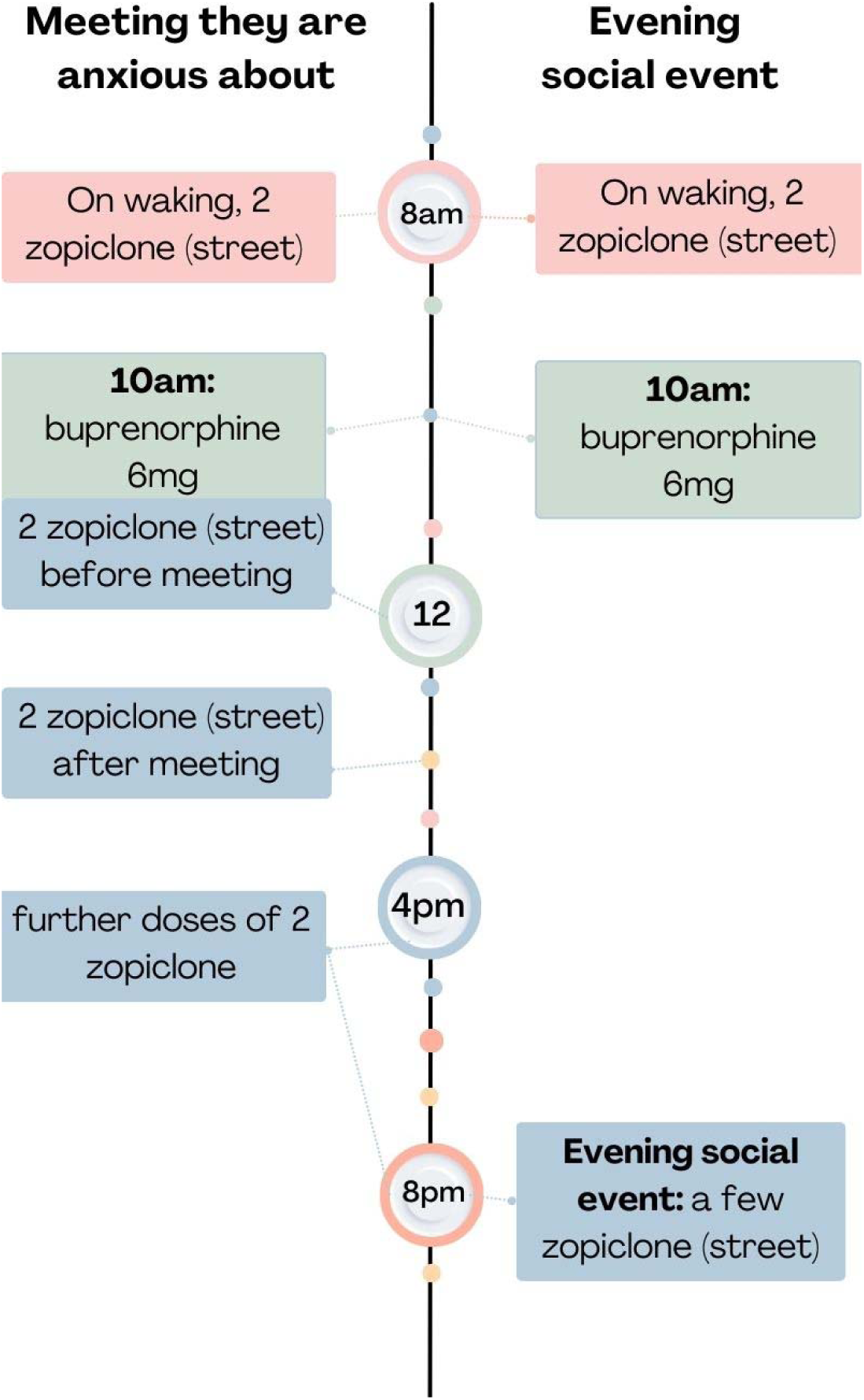
Two co-use timelines as reported by P5 (Male, Teesside), exemplifying how they curated their co-use according to their plans for each day

Both participants had experienced near-fatal overdoses, but only one participant described a near-fatal overdose through co-use, in this instance it was through use of alprazolam and alcohol alongside their buprenorphine.

### 3 Morning and Evening benzodiazepine use with opioids throughout the day

This pattern was characterised by controlled, daily use of benzodiazepines morning and evening, with heavier use of opioids: prescribed methadone (taken in the morning) and illicit heroin throughout the day on top of their OAT. One participant attended heroin assisted treatment (HAT) morning and evening. Compared to pattern 1 and 2, more extensive polydrug use was reported including cannabis (n=2), pregabalin (n=1), cocaine (n=1), crack cocaine (n=1) and alcohol (n=2). It is notable that all the participants reported taking prescribed medicines for their mental health (antidepressants and antipsychotics) which was not a feature of one, but was for one person in pattern two. Two participants had a low-dose benzodiazepine prescription (20-30mg daily dose) and two relied on illicit benzodiazepines taking 10-12 street diazepam or zopiclone split morning and evening. Like pattern two, but not pattern one, participants in this group described co-use as meeting a functional need, taking a low dose of benzodiazepines or z-drugs in the morning before their OAT (e.g. 20mg) helped with anxiety, giving an increased sense of calm and helping to feel like themselves and one further low dose in the evening to help promote sleep:

> “*They make me calm…They just make me feel me.”* (Participant 4, Male, Teesside)

> *“They make me sleep a lot better. Makes me more confident. Makes me a lot better.”* (Participant 6, Female, Bristol)

Figure 3 and the exchange with the interviewer below depicts an example co-use timeline for this pattern, reported by participant 6 (Female, Bristol):

**Interviewer:** So, you take your methadone you say in the morning and when you take your nitrazepam is that at the same time as your morning dose of methadone?
***P6:*** *No. I take it* [nitrazepam] *about 10:00 o’clock in the morning and then I take it about 8:00 o’clock at night*
**Interviewer:** So you take your nitrazepam about 10:00am and…
***P6:*** *Yeah and 8:00pm*.
**Interviewer:** and 8:00am and the methadone what time of day do you take that?
***P6:*** *About 8:00am, 9:00am, as soon as I wake up really*.
**Interviewer:** So you take that first, a couple of hours before.
***P6*:** *Yeah*.
**Interviewer:** Alright and in the morning, when you take your nitrazepam, what does that do for you? How does that help you in the mornings?
***P6:*** *It makes me calm. It makes me think straight. It makes me not shake. It makes me not sweaty.* [note: this participant stated they did not drink alcohol]

Near-fatal overdoses in this pattern were attributed to co-using more drugs than usual. Participant 48, who is prescribed HAT, describes the emotional impact that a low-dose benzodiazepine maintenance script had for him, particularly on near-fatal overdoses:

> *“No. I only started the* [diazepam] *maintenance script when I started the heroin assisted treatment* [HAT]*. When I first got that medication for that, it was very overwhelming. The tears were coming down my eyes, because I’d been waiting for that Valium® maintenance script for a lot, a lot of years, because of the reason being is over the years, I’ve taken that much Valium®, I shouldn’t be here just now. I should be dead. Many times, I’ve been in the hospital with overdoses, and not remembered them.”* (P48, male, Glasgow)

**Figure 3:**
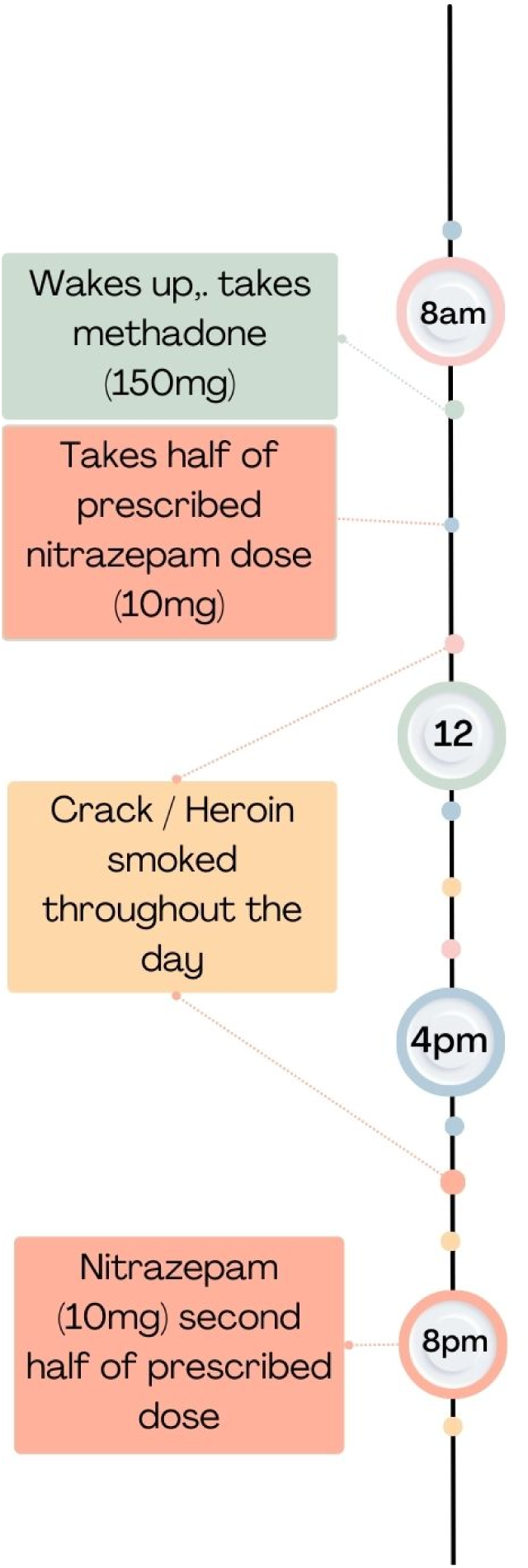
A visual timeline constructed from the descriptions of morning and evening benzodiazepine use with opioids throughout the day as reported by participant 6 (female, Bristol).

### 4 Co-use binges

Co-use binges were characterised by large doses of benzodiazepines or z-drugs and opioids taken over a short time frame (e.g. a few hours) and participants reported that frequently an initial binge would extend to continual binging over a few days. On other days, people relied on prescribed OAT, and often, due to lack of finances following a binge, much smaller amounts of benzodiazepines and illicit opioids were co-used.

Binges typically started through benzodiazepine use, and people described taking *“handfuls”* and eating them like sweets:

> “*I started eating them like Smarties*^1^*, just give me a box of Smarties and I’d just chew them.”* (P9, Male, Bristol).

People in our sample, explained that benzodiazepines reduced their ability to inhibit or stop taking more drugs (opioids, alcohol, benzodiazepines or z-drugs) including describing a sense of invincibility. Others described feeling in control, but heard afterwards from friends or acquaintances that they were ‘out of it’ or had woken up in hospital, prison cells or to destroyed furniture in their home. Co-use binges, with large amounts of benzodiazepine were also reported to cause memory black-outs (see also figure 4 below which depicts the co-use timeline reported by participant 13).

> *“I went through a stage of buying 25 a day as well as the heroin. I would maybe on days I had been paid I would get 100 or 200 and I would wake up the next day with not one left. I couldn’t remember the day before.”* (P13, Glasgow, Male)

**Figure 4:**
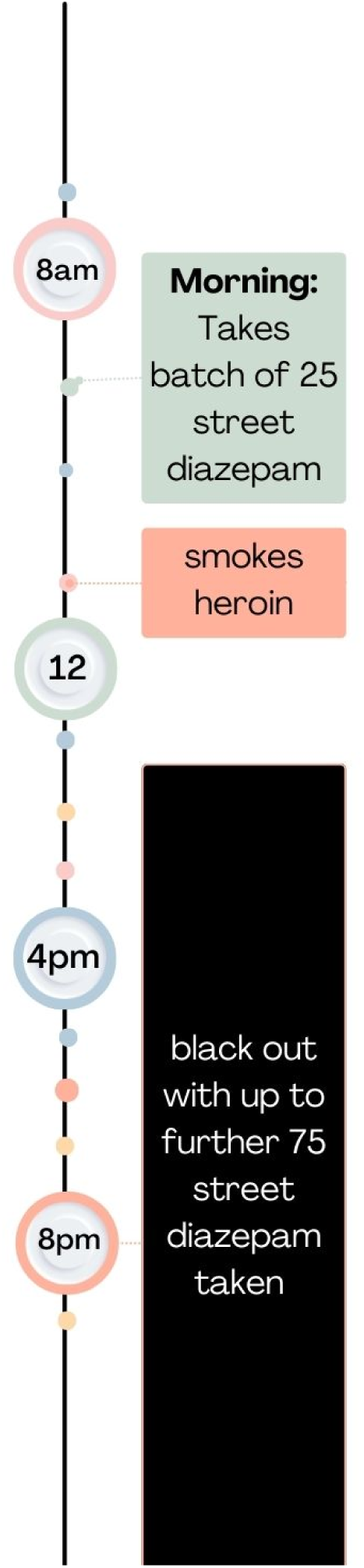
Visual representation of a co-use binge pattern, based on reports from participant 13 (Male, Glasgow)

For some, co-use binges were irregular occurrences with no set frequency or interval between binges (people reported binging on rare occurrences through to 2-3 times per week). However, some binges had predictable timing e.g. once a week when people collected their take home supply of prescribed benzodiazepines and OAT, on pay day, or anniversaries of painful memories such as the birthdays of children taken into care, or loss of a loved one. Co-use binges were also triggered by reminders of traumatic events, or unresolved trauma.

Less predictable (in terms of timing) were binges that were prompted by availability of benzodiazepines e.g. when a trusted dealer had a “good batch” of benzodiazepines. People also reported binges when finances allowed, this was particularly true for those who desired a warm glow, or a buzz from their co-use, finances meant they could chase that feeling by buying large quantities of benzodiazepines. Often, because most people who reported co-use binges were prescribed OAT, the opportunity to access benzodiazepines (due to availability or finances) was the key factor that led to a binge. For participant 23 buying benzodiazepines on pay day led to difficulties managing their drug use:

> “[benzodiazepines] *wouldn’t factor in on just a normal day like that, that’s just my normality taking the methadone but like say if like it’s pay day and I’ve bought benzos or something like that, then everything goes out the window, I couldn’t just take benzos and have my methadone because it would just affecting my brain that makes me want drugs, it would be impossible to stay off drugs if I took benzos. In fact, benzos have made me relapse in the past.* (P23, Male, Teesside)

Binging was part of an intentional overdose for some. Participant 22 describes co-using benzodiazepine and heroin in an attempt to end his life because he was unable to reconcile with himself the unintended harm he caused to someone else whilst taking benzodiazepines:

> *“I never really got over what I’d done* [hurt someone in a fight]*, i regret it every single day but there’s nothing I can do about that now. The trying to overdose thing was just through absolute desperation and the futility of life and all that kind of stuff. Obviously i was still using diazepam. I bought £75 worth of heroin thinking that will definitely, definitely kill me and then I woke up the next day and went ‘bastard’.”* (p22, Male, Glasgow)

### 5 Co-use throughout the day

In this pattern benzodiazepines and opioids (prescribed and illicit) were used throughout the day, re- dosing repeatedly, alongside alcohol and multiple other prescribed and illicit drugs (see Table 4).

Polydrug use was the highest amongst participants in this pattern. Figure 5, and the quote below, depicts a timeline from participant 10 (Male, Glasgow), who reported co-use throughout the day alongside large quantities of crack cocaine and alcohol, whilst at the extreme end of the amounts people reported consuming, similar levels were reported by others who reported co-use throughout the day.

> *I’d get up in the morning and straight away have a drink…half bottle of vodka…put a wee bit of Irn Bru into a bottle, …take either between 25 and 50 street Valium®, out the door down to the chemist. Take 130ml of meth, 30mg of the normal Valium®, already had my pregabalin in the house so they were already taken. From there …depending on how much I had, if I had £200 I could phone up and say, “Give me five rocks or six.” You get three for £50 so that made £100. Then I’d buy them and then I could go to the off licence and get a litre bottle of vodka, down the litre bottle of vodka, another one, litre bottle of vodka. Then buy another litre bottle of vodka and that would leave me with £40 and then I’d maybe go, “Right today I need to buy kit and I need to buy this,” (*P10, Male, Glasgow)

**Figure 5:**
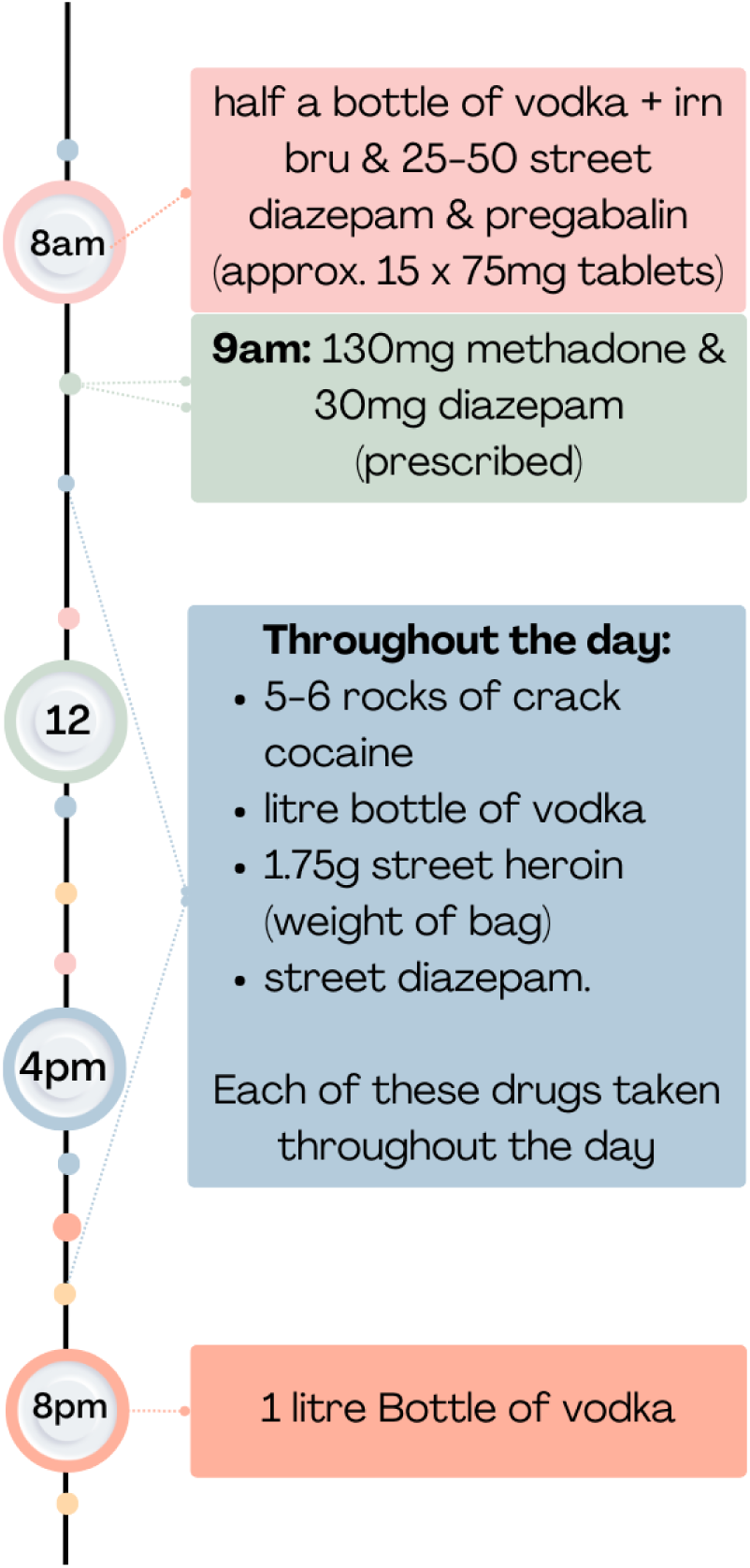
Co-use throughout the day pattern as reported by participant 10 (Male, Glasgow)

**Figure 6:**
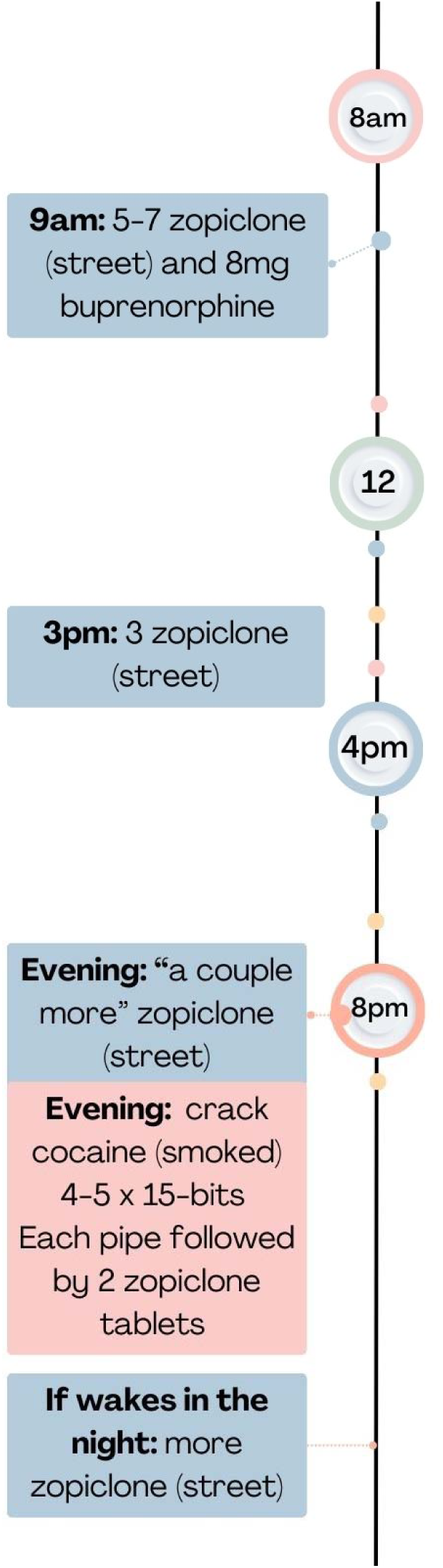
An illustration of participant 18’s z-drugs throughout the day plus OAT timeline

Co-use throughout the day, was linked to chasing the glow or buzz to get to the ‘sweet spot’ and gouching out (typically observed as people slumping over, in and out of consciousness/on the edge of consciousness), or seeking ‘oblivion;’ wanting to be taken away from their surroundings (hostel, homelessness), escape overwhelming mental health symptoms or trauma. Participant 11 describes his daily use of street diazepam described as “street Valium”*®* (alongside prescribed pregabalin, amitriptyline, methadone and human immunodeficiency virus (HIV) medication, and illicit (but not daily) cocaine, heroin and cannabis to reduce re-experiencing past trauma. Below, his quote highlights the ambivalence to the risk of fatal overdose, that many participants who co-used throughout the day reported, such was their need to forget or find a state of oblivion:

> *“I’m in so much pain* [physical and mental] *I just want to forget so I take the Valium® to forget. What I do is I go over the top and that’s why I enjoy them* […] *every time I take them I try and kill myself.”* (P11, Male, Glasgow)

Participant 20 describes her experience trying to hit the ‘sweet spot’ and explained that she had experienced 18 overdoses before she entered treatment:

> *“You get a rush of pins and needles and you’re always chasing that first hit that you never get but you’re chasing it and you’re chasing an overdose but not a lot of people know that.”* (P20, Female, Glasgow)

Like co-use binges, the amounts co-used varied based on availability of drugs or finances. This would make the difference between managing withdrawal symptoms, chasing the glow or seeking oblivion. For some, co-use throughout the day was only about managing withdrawal symptoms and being able to feel well, citing how benzodiazepines gave confidence and reduced anxiety:

> *“It is a case of not what I want, it’s what I need, just to be well. If I’m well, I’m OK. I don’t try and get smashed or oblivion and all that, I’ve gone past all that, I just want to be well.”* (P7, Male, Bristol)

Whilst people acknowledged that their benzodiazepine could affect memory and lead to disinhibited behaviour, memory black-outs were not frequently reported but was accepted that this was a consequence or desired outcome of their co-use. As the quotes from participant 11 and 20 above exemplify, people were also ambivalent about the overdose risk posed by their co-use throughout the day.

### 6 Benzodiazepines or z-drugs throughout the day plus OAT

We distinguished this pattern from co-use throughout the day, because opioid use was limited only to prescribed OAT. Often, this was because people could no longer successfully inject, or were receiving long-acting injectable buprenorphine and therefore heroin was perceived to be a waste of money, although some participants described taking an occasional dose of heroin to test if things had changed. In place of illicit opioids, participants described how their benzodiazepine/z-drug use had increased once stabilising on injectable buprenorphine or once they realised they could no longer inject. Others described intentionally avoiding illicit opioids to reduce the risks of their benzodiazepine use, choosing other drugs (e.g. cannabis) to use alongside their benzodiazepines and OAT.

> *“Cannabis* […] *is the safest of them, it can’t hurt you.”* (P35, Male, Glasgow)

Large amounts of benzodiazepines (e.g. 100 street diazepam) were taken throughout the day, re- dosing, alongside other illicit drugs – often crack cocaine or cocaine and prescribed medicines. Below a series of quotes, and a visual depiction of Participant 18’s co-use timeline is presented.

> *“I wake up, as soon as I wake up, between 5 and 7 and then that will do me until about, just say it’s 9 o’clock I take them* [zopiclone]. *That will do me until about maybe 3, 4 and then I’ll neck another 3 and then I’m necking a couple more to go to sleep and if I wake up in the middle of the night, I’m necking more…Half an hour after it* [the 5 zopiclone at 9am in the morning]*, about half an hour. I have a couple with my Subbies under my tongue so I’ll have a cuppa, neck the tablets…The other night I have five 15s, like 15 pound bits* [£15 rocks of crack cocaine]*. The night before I had four but then as soon as I have pipe I’m necking two tablets* [zopiclone].”

Behavioural routines had been developed through trial and error that enabled people to prevent a prolonged binge pattern and avoid the immediate harms caused by high benzodiazepine or z-drug use (e.g. getting into trouble, memory blackouts). These routines included only co-using at home, alone (to avoid being taken advantage of, or their disinhibited behaviour leading to conflicts), leaving notes to themselves about where their keys, phone, wallet were before they started taking drugs. Near-fatal overdoses were experienced with this co-use pattern, but participants reported wanting to avoid a fatal overdose, rather than ambivalence, and described them as *“accidental”* or *“took too many.”*

> *“Accidental, twice. That’s the hardest part I don’t get about it, is it was accidental but it was twice it happened in the space of two days.”* (P34, Male, Teesside)

### 2. Overdose risk and co-use patterns

Table 5 reports the numbers of people in each pattern who had ever experienced a near-fatal overdose^2^ through drug use, and of these how many of these they attributed to their current benzodiazepine opioid co-use pattern. For all patterns, at least one participant reported a near-fatal overdose and a trend is observed, where a higher proportion of participants prescribed methadone reported near-fatal overdoses compared to participants prescribed buprenorphine. As described above, not all co-use patterns were daily patterns, and not all patterns involved high doses of benzodiazepines and/or opioids which may give the sense of a lower risk of fatal overdose compared to daily, less controlled patterns of co-use (patterns 5-6). However, as highlighted at the end of each pattern, non-fatal overdoses did occur. Potential explanations for this are because of an unexpectedly strong batch of benzodiazepines/z-drugs or because they had temporarily switched into a less controlled pattern of co-use. It’s important to note that people were also aware that benzodiazepines and opioids could be a fatal combination, and specifically combined these drugs to intentionally overdose.

**Table 5:** Numbers of near-fatal^a^ overdose experiences ever and near-fatal overdose experiences through current co-use pattern and opioid agonist therapy (OAT) taken.

|  | **No pattern** | **Co-use to aid** | **Curated co-use** | **Morning and** | **Co-use binges** | **Co-use** | **BZ/Z use** |
| --- | --- | --- | --- | --- | --- | --- | --- |

|  | reported<sup>b</sup> (n=5) | sleep or<br>come down<br>(n=3) | (n=2) | evening BZ/Z<br>doses with OP<br>throughout the<br>day (n=4) | (n=13) | throughout the<br>day (n=14) | throughout the<br>day, OAT only no<br>other OP (n=7) |
| --- | --- | --- | --- | --- | --- | --- | --- |
| Number of<br>participants who had<br>ever had a near-fatal<br>overdose experience<sup>a</sup> | 3<br>(OAT prescribed at<br>time of this OD<br>methadone n=2,<br>buprenorphine<br>n=1) | 2 | 2 | 2 | 11 | 12 | 5 |
| Number of<br>participants who had<br>a near-fatal overdose<sup>a</sup><br>using their <u>current</u><br>co-use pattern (OAT<br>prescribed at time of<br>this OD) | n/a | 1<br>(methadone<br>n=1) | 1<br>(buprenorphine<br>n=1) | 2<br>(methadone<br>n=2) | 9<br>(methadone,<br>n=6,<br>buprenorphine<br>n=1) | 9<br>(methadone,<br>n=5,<br>buprenorphine<br>n=1) | 5<br>(methadone, n=3,<br>buprenorphine<br>n=2) |

<sup>a</sup> Near-fatal overdose defined as receiving treatment from a paramedic, police officer, friend or family member (e.g. resuscitation, naloxone rescue) and/or hospital admission for overdose. It does not include events where people report prolonged periods of unconsciousness without intervention.
<sup>a</sup> Some participants could not be mapped to a pattern because their interview did not capture sufficient information on when they took their opioids and benzodiazepines

For most people in our study these drugs were used in combination with a range of prescribed (e.g. antidepressants – mirtazapine, anti-inflammatory pain medication – naproxen, anticonvulsants- pregabalin) (see table 3). These medications may also interact with the benzodiazepines and opioids they are taking. It can also be seen in table 3 that people were taking a range of other illicit drugs - crack cocaine, street/diverted pregabalin, cannabis, spice and drinking alcohol which also could affect the risk of overdose whilst co-using. Some participants perceived additional risks of drinking alcohol whilst co-using, however, the risks of different combinations of drugs on-top of their benzodiazepines and opioids were not well understood, beyond the sense that more drugs equated to more risk.

Following a near-fatal overdose, some people described a change in pattern to reduce their risk of future overdose. However, all participants described how their co-use patterns changed over time, reflecting either an escalation or de-escalation in their drug use, describing that drug use escalated as tolerance to the euphoric effects of benzodiazepines occurred, or because their circumstances or mental health worsened, leading to drug use to forget or seek oblivion. Patterns also changed, following a switch in methods of administration of opioids (e.g. from injecting to smoking heroin), or in response to availability and/or access to prescribed or illicit drugs.

### 3. Order in which opioids and benzodiazepines were taken

There was no clear preference for taking one drug before the other. However, many participants reported trying to save a few of their benzodiazepines for the next morning. The motivation for this was functional, for example, to help them get out of bed, describing their first benzodiazepine dose to help give them energy and confidence to leave the house or to prevent benzodiazepine withdrawal effects which participants perceived to be worse than opioid withdrawal effects. Other participants reported taking opioids first, and then adding in benzodiazepines or z-drugs either to “bring on” or sustain the effects of an opioid.

### 4. Spread of participants across patterns in relation to prescribed OAT/benzodiazepines and risk of overdose

The relationship between the type of OAT prescribed and patterns of co-use appears complex. On the one hand each pattern (except curated co-use) included participants prescribed different types of OAT (see Table 4) suggesting patterns were not related to prescribed OAT. On the other hand, a change in prescribed OAT (e.g. switching to long-acting injectable buprenorphine) or the amount of OAT was reported to trigger a change in co-use pattern. A change in prescribed benzodiazepine could trigger a change in co-use pattern too. Participant 8 describes the impact the discontinuation of their long-term low-dose benzodiazepine prescription had on their co-use pattern:

> *“I had a problem with benzos for years, like buying them on the street and I’m buying them online as well. I’ve been with various GP surgeries, I was prescribed diazepam for quite a few years and that then kind of held me fine, I was absolutely fine, and then the GP surgery I’m with now, about two years ago, they wanted me to start chipping away at the dose I was on, and I didn’t feel I could say no because of the medication. I felt that if I said no I’m not ready to do it then they would have just done it anyway, so they chipped away 2mg every month or so from 30mg all the way down. It was last year in about spring time that I actually came off of it completely and within a couple of weeks of that happening I was buying massive amounts online and literally going into overdose mode. I was taking 60 in one two day session.”* (P8, male, Bristol)

Some participants felt that a benzodiazepine maintenance prescription would stop their need to buy illicit diazepam and give them access to safer benzodiazepines than their street supply. Whilst others who had benzodiazepine prescriptions reported taking their prescription over a couple of days and then buying street or diverted prescription benzodiazepines to cover the rest of the week. One participant who had an OAT prescription stated: *“No, one maintenance prescription is enough”* (Participant 30, Glasgow, male) when asked if benzodiazepine maintenance prescribing could be helpful.

### 5. Demographic/ socioeconomic spread of participants across patterns

Each pattern was reported by male and female participants and across all study locations, with the exception of curated co-use which was only reported by two participants. One contextual finding between the three areas was people’s reports of local drug markets. Glasgow participants explained all benzodiazepines sold on the street were sold as “street Valium®” and that people knew this could be any type of benzodiazepine, but not which one, limiting their ability to be selective.

> *You had no idea what’s in them* [street Valium®] *and that’s why there’s three deaths a day or something like that in Glasgow just now. That’s what scared me and made me start paying through the nose to get the real ones so I at least knew what I was getting.”* (P22, Glasgow, Male)

Bristol participants reported that a range of illicit benzodiazepines were available locally, as well as diverted prescription benzodiazepines, and many sourced them online, which enabled curated co-use (or at least a perception of curated co-use). In Teesside, people reported illicit or diverted benzodiazepines were hard to find but illicit zopiclone was readily available. No participants in Teesside currently held a benzodiazepine or zopiclone prescription, whereas 17 participants amongst those recruited from Bristol and Glasgow did. Seven participants from Glasgow were prescribed the buprenorphine long acting injection but no participants from Bristol or Teesside were. Most participants recruited in Glasgow reported current co-use patterns that sit in the less controlled category (co-use throughout the day, benzodiazepine/z-drug use throughout the day plus OAT, or co-use binges).

## Discussion

We propose a continuum of co-use overdose risk, featuring six patterns of co-use which future population level studies could quantitatively validate.

Previous studies have quantitatively described co-use patterns (24, 43–46). Latent class analysis has identified three types of co-use: 1) benzodiazepine, cocaine and alcohol use; 2) former heroin, cocaine, tetrahydrocannabinol and alcohol use and 3) benzodiazepine only (45). Our study builds on this work, providing details on patterns of co-use: the times of day each drug is taken, reporting how and when each drug is taken in relation to each other, and for what reason. We have also distinguished how prescribed and illicit opioids and benzodiazepines/z-drugs are co-used in each pattern, and highlighted the wide spectrum of polydrug use, and other prescribed psychoactive medication taken alongside. This study has also highlighted two previously unreported patterns of co-use; benzodiazepine/z-drug use throughout the day plus OAT and curated co-use. In both these patterns, people reported using very little illicit opioids, and being stabilised on their OAT, but a greater reliance on benzodiazepines.

The approximate number of benzodiazepine tablets taken in our study is similar to previous research by Ross et al (43) which reported a median of three tablets but a wide range of 1-150 tablets, but did not report the strength or content of the tablets taken. In our study, we found that co-use binges, co-use throughout the day, and benzodiazepine/z-drug use throughout the day plus OAT, were characterised by higher overall daily intake of benzodiazepine / z-drug tablets compared to the other three patterns. Furthermore, most of our interview participants co-used daily, but those who reported binges or using benzodiazepines / z-drugs to sleep or come down reported less regular co-use. In contrast, a retrospective 7-day diary study found only 10% of participants reported same day use of opioids, benzodiazepines and alcohol in the past week, two-thirds used opioids daily in the past week, and more than half reported at least one day in which they used all three substances (44). This suggests our interview sample were potentially more entrenched in their co-use, alternatively, OAT dosing did affect switches in co-use patterns, and it is possible that perceived inadequacy of OAT dose may influence, a greater reliance on benzodiazepines and other drugs.

People taking OAT in Baltimore and Philadelphia in the US reported using 40-45mg (median) diazepam the majority (82% of Philadelphia participants, and 92% of Baltimore participants) taking it once daily within an hour of using their OAT medication (24) citing that diazepam enhances the effects of methadone, which has been supported elsewhere (47). This differs from the patterns reported here, where most patterns included people who took benzodiazepines more than once per day. However, this difference might reflect changes in the illicit benzodiazepine and opioids available, and prescribing patterns over the last 40 years. Vogel et al. (46) report over half their participants who co-used divided their benzodiazepine dose – this was more common among participants with high negative affect regulation (e.g. “to lose anxiety” or “to forget problems”). This aligns with our current study’s co-use patterns: morning and evening doses and curated co-use with explicit purposes of managing anxiety and poor sleep. The motivation for splitting benzodiazepine dose “to feel better in the morning” and “to improve sleep” at night identified by Vogel et al. (46), also mirrors our findings in the morning and evening doses of benzodiazepine plus opioid use throughout the day pattern. Stemming from this there are a number of interventions that could be tailored to co-use patterns that involve a low dose of benzodiazepine once a day or split into two doses. For example low intensity psychosocial interventions could include, emotion regulation skills (e.g. as taught in dialectical behaviour therapy) or anxiety coping skills to aid with reducing anxiety in the morning and evening. Such approaches may allow people to reduce their reliance on benzodiazepines to address their negative affect. Alternatively, prescribed antidepressants may be a solution, although a number of our participants were already prescribed antidepressants and antipsychotics. More intensive psychological therapy is also urgently needed, however people receiving substance misuse treatment in the UK often fall in the gaps between substance misuse and mental health services (48)

It is notable that participants in the present study reported co-use as a method of coming down from stimulant use in pattern 1, and a number of participants in patterns 4-6 reported use of crack cocaine. Stimulant use, and fatal overdoses involving stimulants have increased sharply in recent years in the UK (11, 49). Stimulant use in addition to benzodiazepines and opioids may further increase the risk of fatal overdose.

Co-use can vary from using benzodiazepines prior to, after or at the same time as heroin and OAT depending on the motivation for use (23, 24, 27). In agreement with previous research the current study found that the order in which opioids and benzodiazepines or z-drugs were taken, as well as the timing, frequency and dose could be explained by the motivations underpinning the pattern. For instance, in pattern 2, curated co-use, the timing and dosing was varied to achieve a specific function (enhance confidence, reduce social anxiety) or achieve a euphoric state (warm glow or buzz), in pattern 3, benzodiazepines were taken first, to manage symptoms of anxiety so that they were then able to get out of bed and start their day, with opioid use happening later compared to co-use throughout the day where large doses were taken throughout the day motivated by achieving a buzz or glow or seeking oblivion in response to mental health symptoms or trauma. These relate to a set of ‘functional’ and ‘experiential’ motives for co-use identified in the data, reported in more detail elsewhere (32). To some extent these motivations reflect previously identified overarching drivers of co-use: 1) self-therapeutic (removing negative emotional states such as anxiety) and 2) hedonic (or euphoria seeking) (46, 50, 51). Furthermore, frequency of benzodiazepine use among people with OUD has previously been associated with higher levels of anxiety sensitivity (52).

The opportunity to co-use benzodiazepines supported by their accessibility, availability and finances (13, 53) facilitated the binge pattern of co-use in our sample. In contrast, the curated co-use pattern points to considerable deliberation about each day’s co-use. Similarly, Chang et al, found that polysubstance use is characterised by a ‘delicate balance, planning, and calculation’ (p30) (54).

Co-use patterns may be influenced by location, as participants in Glasgow reported following the patterns that represented less controlled co-use (co-use binges, co-use throughout the day and benzodiazepine/z-drug use throughout the day plus OAT). Given the higher levels of drug related deaths in Scotland and particularly Glasgow this finding is not unexpected. However, to ensure this finding was not a reflection of the sites that participants were recruited from in Glasgow, a residential stabilisation unit and a crisis service, recruitment was expanded to other services providing care and harm reduction support in the community, both within the city centre and in a smaller town outside Glasgow. At the same time, in Bristol, we worked with peer researchers who supported recruitment of participants who were not currently engaging with treatment and/or living away from the city centre to see if this enabled us to capture experiences similar to those reported in Glasgow. These efforts expanded the range of experiences reported in our study and participants reporting more controlled co-use in Glasgow, but we also continued to find evidence of less controlled co-use amongst participants receiving treatment in the community.

The present study’s findings have implications for policy and practice related to co-prescribing and wider harm reduction efforts to address DRD among people who co-use. Understanding patterns of co-use and how they vary may help identify people who would benefit from co-prescribing or not. Participants had mixed views about benzodiazepine maintenance prescriptions despite promising evidence of acceptability reported by research conducted in Scotland (55). The benefits of accessing a safer supply were offset by insufficient dosing necessitating additional use of street or diverted benzodiazepines on top. Indeed, discontinuation of prescriptions without shared decision making with patients can risk disengagement and use of illicit street benzodiazepines (56), this was also reported in our study. Co-prescription of opioids and benzodiazepines is associated with an increased risk of death (4, 16, 17, 57, 58), though it is also associated with OAT retention. Prescription of benzodiazepines is one of the most consistent correlates of ‘misuse’ (use without a prescription; at a higher frequency or dose than prescribed) (57, 59, 60). Taking prescribed benzodiazepines in ways other than as prescribed may be motivated therapeutically (60–63) or intentionally (13, 43, 60, 64). As a result, benzodiazepine prescribing practice is recommended to include information about risks, safe medication storage and disposal to help prevent diversion and accidental exposure (60). While patients are aware of the risks of co-use, they continue to use benzodiazepines with OAT for the benefits (56). In contrast, clinicians focus on the risks over the benefits which can result in differences in treatment goals (discontinuation for clinicians and maintenance or long-term use for patients)(56). Advice has warned against co-prescription of benzodiazepines and opioids for people who are likely to binge use (65), however participants in our study reported shifting to a binge pattern when low dose benzodiazepine maintenance prescriptions were discontinued. Further guidance from the Medicines and Healthcare products Regulatory Agency (MHRA) has advised against co-prescribing (66) due to the overdose risk. The concerns of clinicians need to be explored further in conjunction with the wishes of people who co-use.

There is variation in the practice of co-prescribing. In Scotland, a survey of 55 prescribers, found that two-thirds were prescribing benzodiazepines for people who use opioids and were dependent on benzodiazepines and ∼80% would be willing to prescribe. Those who did not prescribe cited insufficient evidence, local guidance, organisational preferences for patient self-detoxification, difficulties monitoring on top use and potential harms leading to drug related death (53). A systematic review of qualitative studies highlighted ambivalence among GPs towards prescribing benzodiazepines (not in relation to opioid co-use) and inconsistently applied strategies for managing their use. Prescribing decision making was ‘often uncomfortable, demanding and complex within the time and pressure constraints of daily practice’. Tensions between helping patients and minimising benzodiazepine prescribing were managed on an individual basis (67). Such patient-centred treatment could support prescribing decision making. A commissioned call has been made for randomised controlled trials to assess the risks and benefits of co-prescription of benzodiazepines for people in opioid agonist treatment and inform clinical guidance (57).

Our findings, also suggest that a change in prescribed OAT, benzodiazepines or other psychoactive medications triggered a change in co-use pattern. Such changes could represent a critical window in which to monitor health and overdose risk. Routine, systematic measurement of the patterns and motivations for co-use, using tools like an Inventory of Drug Taking Situations (68) could help support people with tailored advice and treatment and contribute to the consideration of prescribing decision making (53, 69). Most participants in our study reported use of benzodiazepines / z-drugs alongside opioids to manage untreated mental health symptoms and trauma, and patterns varied depending on the types of symptoms and trauma they were trying to self-medicate. Treatment of mental health symptoms such as anxiety could also help address the therapeutic uses of benzodiazepine alongside opioids (52). Previous qualitative evidence captured a process of people on OAT learning to use benzodiazepines in a safe, therapeutic way (56).

Heart rate and respiratory depression, caused by co-use, increases the risk of overdose (26). In this study, non-fatal overdoses were mostly reported by those reporting less controlled co-use (patterns 4- 6), pointing to a hypothesis that there may be different levels of overdose risk related to patterns of co-use. One plausible explanation for infrequent binges increasing the risk of overdose is that tolerance to the drug’s effects may remain low (44). Awareness of overdose risk associated with co-use varies among people who co-use (70–74). Participants who co-used throughout the day, reported co-using alone, to avoid being taken advantage of, or getting into conflicts because co-use caused disinhibited behaviour. However this strategy runs counter to traditional harm reduction advice to take drugs around others so that help is on hand if needed. Alternative strategies may be needed, for example research is underway in a number of countries into the potential for wearable devices to detect respiratory depression (75), and the potential for the wearable to administer naloxone to reverse the effects of opioids taken (76) although these devices may not be acceptable to all, particularly if people were seeking a state of oblivion. For those seeking oblivion, drug consumption rooms may provide a safe place for people to co-use around others, without being vulnerable to the consequences of disinhibited behaviour or being taken advantage of. In addition to harm reduction messages to raise awareness of these risks among people who co-use and continued provision of naloxone (44), scale up of drug checking services, drug consumption rooms and co-prescribing for people who co-use needs to be explored (77). Drug checking services could hypothetically better help people who co-use, through providing information on the contents, strength and potential dose in their illicit drugs and quantifying the risk of combining these substances alongside any other prescribed medication. A recent evaluation of a community based drug checking service highlighted the benefits that drug checking could bring for people who co-use. On receiving information about the contents of their drugs participants reported intentions to take greater car e mixing that substance, taking less of the substance(s) and alerting friends to the contents (78).

### Findings related to the socioecological model (79) and co-use

At the **intrapersonal level** our findings highlight that people co-use benzodiazepines or z-drugs to self-medicate mental (anxiety, depression, trauma) and physical health symptoms (withdrawal, pain) or achieve specific states of euphoria (warm glow/buzz) or oblivion. We have also highlighted that individual’s knowledge of, and concern with overdose risks varies. Individual finances were also a factor that affected co-use. At the **interpersonal level** the study findings suggest that co-use is also achieves the function of increasing confidence for people who experience social anxiety. Some participants reported the availability of drugs was a trigger to co-use – for example participants reported co-using following an offer of illicit opioids, benzodiazepines or z-drugs by a friend, acquaintance or dealer – particularly if it was their preferred type. Conflict, relationship issues, abuse and trauma imparted on them or by them was also a trigger for uncontrolled patterns of co-use as a way of dealing with these challenges. At the **organisational level** OAT and benzodiazepine or z-drug prescribing decisions impacted on people’s patterns of co-use. At the **community level** it was evident that local drug markets had an effect on patterns of co-use both in terms of the types of illicit benzodiazepines or z-drugs taken and the numbers of people reporting less controlled co-use patterns. Findings at the **policy level** are reported elsewhere (33, 40).

### Strengths and limitations of the work

In the absence of drug testing, the study relied on self-reported substance use, which means, the illicit benzodiazepines / z-drugs and opioids people report using may have contained other substances and doses reported may have been inaccurate. Some participants experienced difficulties recalling the specific timing, strength and number of doses. However, collecting this data through interviews meant that researchers could probe for depth of details and context that people may not have thought to disclose in a questionnaire or diary study. Details of co-use prior to a near-fatal overdose was challenging for some participants to recall. However, the insights generated are still important as behavioural responses following these experiences contribute to the risk of future DRD.

The broad range of co-use experiences captured in three localities, which each have different drug markets, drug and alcohol treatment policies, services and DRD rates is a major strength of this work. A further strength is the input of people with lived experience to the development of the research idea, conduct of the interviews and interpretation of the findings.

## Conclusions

The patterns identified provide opportunities for future harm reduction strategies, tailoring advice to patterns of use, updated prescribing guidance and policies, and the need for better access to mental health care, for people who co-use benzodiazepines and opioids to reduce DRDs.

## Data Availability

Data are available on application at the University of Bristol data repository data.bris. Data access is restricted to bona fide researchers for ethically approved research and subject to approval by the University's Data Access Committee.

https://data.bris.ac.uk/data/

## Consent for publication

All interview participants provided written informed consent to participate in the study, and consent for anonymised quotes from their interviews to be included in published work.

## Availability of data and materials

Data are available on application at the University of Bristol data repository, data.bris.ac.uk. Data access is restricted to bona fide researchers for ethically approved research and subject to approval by the University’s Data Access Committee. An abbreviated topic guide for the study is included in appendix

## Authors’ contributions

**HF:** methodology, validation, formal analysis, investigation, data curation, writing – original draft, writing – review and editing, visualization. **GV& HP:** methodology, validation, investigation, data curation, writing – review and editing. **MH, CB & APAS:** Funding acquisition, conceptualization, validation, writing – review & editing. **GH:** Funding acquisition, conceptualization, methodology, validation, writing – review & editing, supervision, project administration. **JK:** Funding acquisition, conceptualization, methodology, validation, writing – original draft, writing – review & editing, supervision, project administration, data curation. **JS:** Methodology, validation, writing – review & editing, supervision, project administration, data curation and clinical interpretation. **SK, DC & SS:** validation, writing – review & editing. **NB, PDS, LA:** Investigation, validation, writing – review & editing.

## Acknowledgements

We would like to thank the research participants for sharing their experiences with us and the services and staff that worked with us in Bristol, Teesside and Glasgow. We would also like to thank the peer co-researchers, Joanna Green, Chris Shilvock, Jade Ritchie, Louise Aitken, Nick Booth and Peter Da Silva. Thanks also to the expert stakeholders who have acted as critical friends, reviewed the findings and given their feedback on the relevance for future policy and practice. The views expressed are those of the authors and not necessarily those of the Medical Research Council, the NIHR, the Department of Health and Social Care, or UKHSA.

## List of Abbreviations

UK: United Kingdom
DRD: Drug Related Death
OAT: Opioid Agonist Therapy
OUD: Opioid Use Disorder
GABA_A_: g-aminobutyric acid type A receptors
CLIP-Q: collaborative and intensive pragmatic qualitative
HAT: Heroin Assisted Treatment
HIV: Human immunodeficiency virus

## Appendix: Interview topic guide

|  |  |  |
| --- | --- | --- |
| **1. Introduction** | Explain purpose of research project | *The purpose of this study is to understand the experiences of people who use benzo and opioids. You are the expert and I really appreciate you sharing your views with me.* |
|  | Explain audio recording procedures | *Before we get started, I'd like to tell you that I will be recording the conversation to help us remember what we discussed. You can ask for the recording to be stopped at any time. What you say will be kept confidential and anonymous.* |
|  | Check documents complete and participant has opportunity to answer questions | ➤ Check participant is happy with peer researcher being present<br>➤ Go through differences in peer researchers role to main role (if applicable) and divert any requests for advice and support to after the |
|  |  | interview (if necessary).<br>➤ Ensure participant has copy of participant information sheet<br>➤ Answer any questions<br>➤ Explain that you will ask about overdose experiences and check they are comfortable with that topic?<br>➤ Complete consent form [written or verbal (if telephone interview) – read out statements as needed]<br>➤ Commence audio recording |

## 2. BACKGROUND INFO

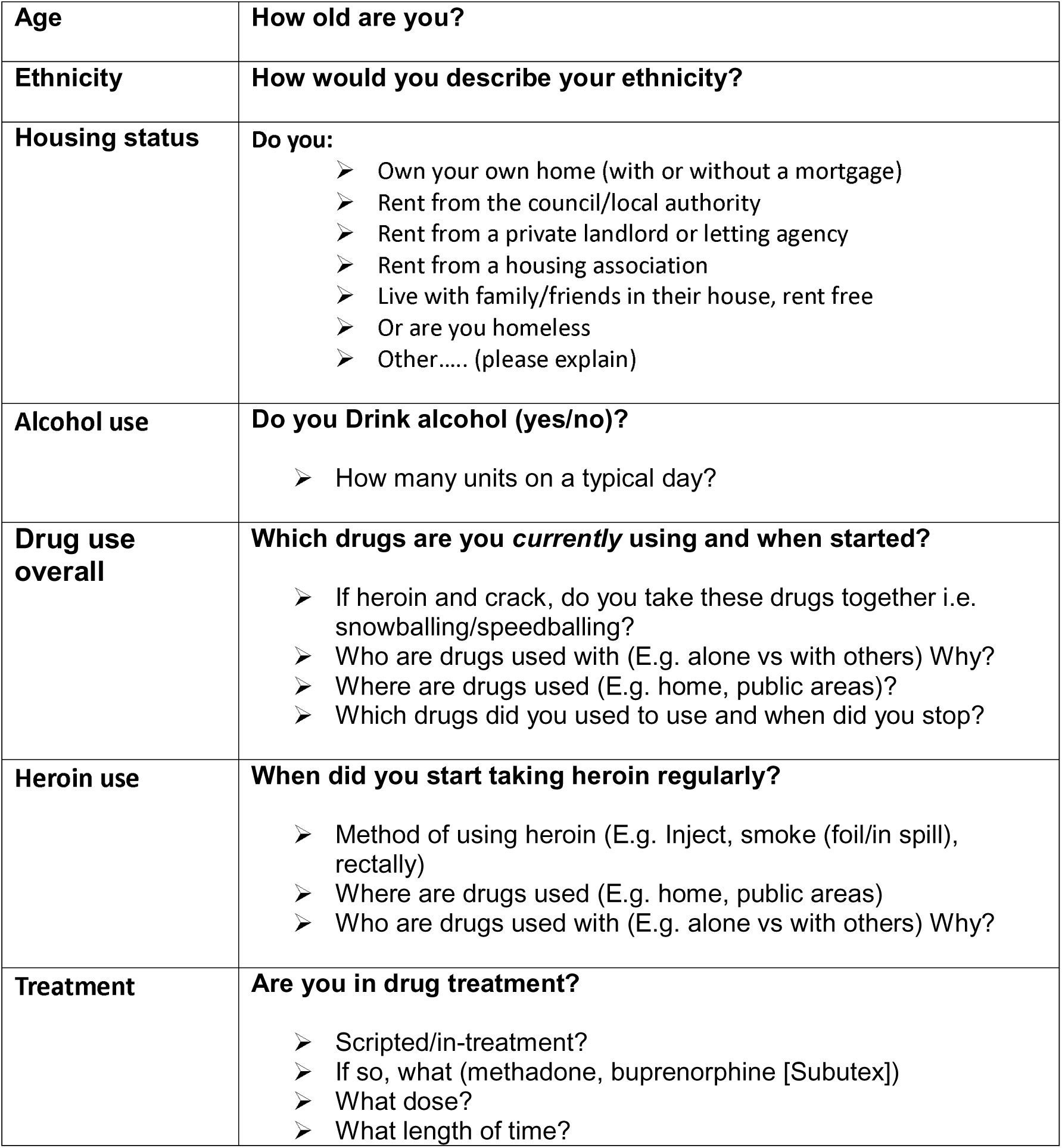
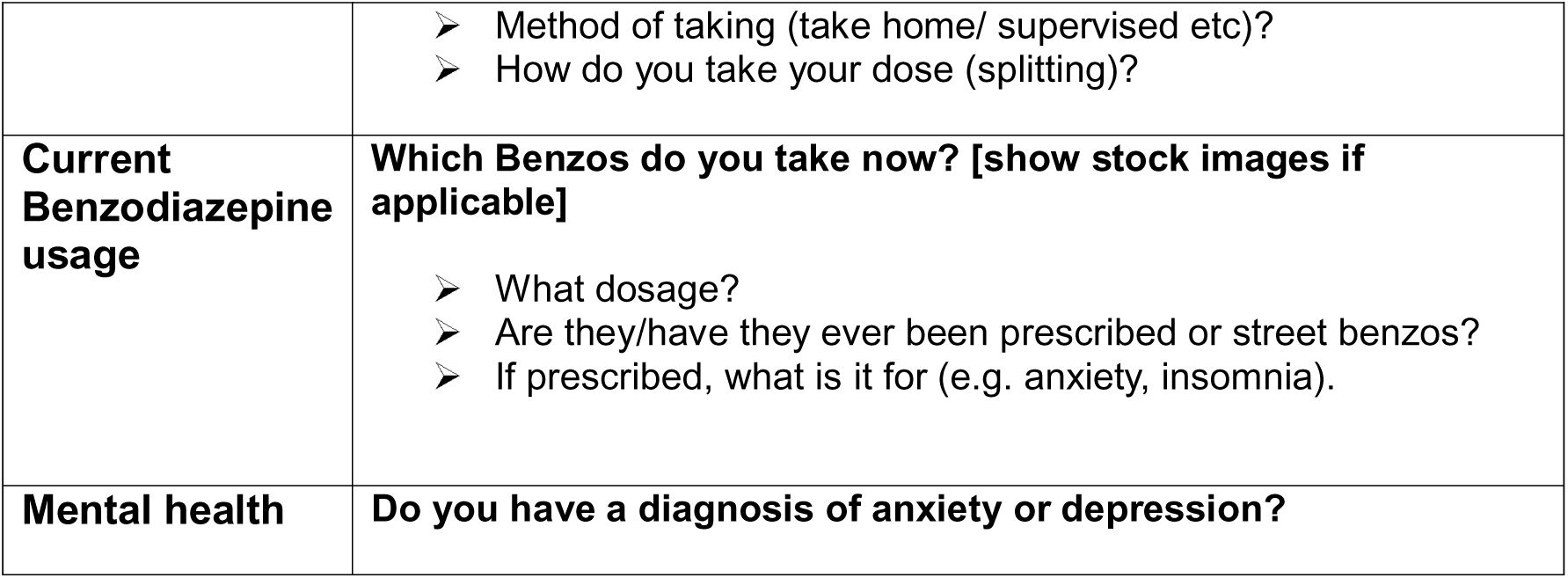

## Main body of interview que

Ask questions in bold first, follow-up with questions underneath as needed

|  |  |  |
| --- | --- | --- |
| 3. | **How did you start BZ's alongside opioids** | **When did you started using benzos and why?**<br>**How started using benzos AND heroin or a script?**<br>➤ Which drug started first, why, or always used BZs and opioids?<br>➤ Ask about benzo / opioid co-use in the context of detoxing (either by themselves or with a detox service)<br>N.B. Make sure you're clear about type opioid (i.e. is it methadone, Subutex (buprenorphine), Buvidal (implant), or heroin) |
| 4. | Current method/experience of using BZs and opioids | **HAVE IMAGES OF ZOPICLONE/STREET BENZOS TO HAND TO AID IDENTIFICATION**<br>**Do you have a preferred type of Benzo? i.e. imagine if there was a place or a shop you could get ANY benzo or z-drug which would you choose?**<br>➤ why would you choose this benzo/zdrug?<br>➤ What is good about this benzo/zdrug?<br>➤ [probe: is etizolam preferred to temazepam for example?]<br>➤ Where are BZs accessed from?<br>➤ How much do they cost?<br>➤ How do you know they are what they are sold as? For example, zopiclone (e.g. 'metal mouth'/ taste test for zopiclone users)<br>**Describe a typical day in terms of how you use BZs and opioids? Please give as much detail as** |
|  |  | **you can.**<br>➤ Which substance used first or are they taken at the same time/situation and why?<br>➤ What time do you take each substance? Why is that?<br>➤ How often do you re-dose (e.g., how long do the effects of different BZs last)?<br>➤ What leads to re-dosing (e.g. tolerance/dependence/ sleep)?<br>➤ How much BZ is taken (including the practice of 'mega-dosing' (using handfuls))?<br>➤ Do you use benzos differently when using heroin, methadone or buprenorphine? (amount, frequency of re-dosing)<br>➤ Do you take benzos before you go to sleep?<br>**What are your reasons for these patterns of use?**<br>➤ E.g. sleep, poor mental health, physical dependence, habit<br>➤ If physical dependence, explore whether it is opioid or bz dependence – do they know which it is? If yes why, if no, why not?<br>**How has your use of BZ and opioids changed over time?**<br>➤ How did you used to use them?<br>➤ Why has this changed?<br>➤ What is your personal experience of tolerance to BZs and opioids? Does tolerance influence how you use these drugs? Probe around any detox attempts (self-detox or in clinic) and whether tolerance changes after detox and if so their experience<br>**What do you like about using BZs and opioids?**<br>Prompts: sedative; like the feeling; long-lasting; remove withdrawal symptoms; enhance opioid effect; self-medication for mental health problems, insomnia, pain or injury, difficulties accessing prescribed medication; poor quality heroin; experiences of stigma preventing healthcare access; and suppressing harmful side effects associated with stimulant use; counteract effect of |
|  |  | stimulants<br>**What do you dislike about using BZs?**<br><u>Prompts:</u> memory loss including losing belongings, cognitive impairment and blackouts<br>Definition: Cognitive Impairment is the medical term for problems with any aspect of thinking ability.<br>Symptoms of a Cognitive Impairment can include:<br>➤ Memory problems<br>➤ Understanding<br>➤ Reasoning<br>➤ Planning<br>➤ Problem solving<br>➤ Decision making<br>➤ Attention<br>➤ Difficulty interpreting the world around (visual-spatial skills)<br>➤ Speech and language (e.g. word finding difficulties) |
| 5. | **Experience and perceptions of overdose risk** | **Is overdose something you think about specifically with benzos & heroin/methadone/bupe?**<br>➤ Do you worry about it?<br>**Have you had a personal experience of overdose when using benzos & heroin/methadone/bupe?**<br>➤ How recent was this?<br>➤ Do you think the risk of overdose differs with different type of BZs? Are any specific benzos more dangerous than others?<br>➤ Does the risk differ with different types of opioid – heroin/methadone/buprenorphine?<br>➤ Do you think there is a greater risk of overdose when taking benzos and heroin/methadone/bupe over just heroin/methadone/bupe opioids alone?<br>➤ What influences these risks? E.g. going to sleep, lying down / slumping forward<br>**Can you describe the most recent or a couple of experiences of overdose when using BZs and opioid?**<br>➤ How did this feel? |
|  |  | ➤ Notice breathing affected?<br>➤ What drug were you taking at the overdose?<br>➤ What do you think happened/ why did you overdose?<br>➤ Did someone use naloxone to revive you? How did this feel? Or are you unsure?<br>➤ Did you experience memory loss, cognitive impairment and blackouts caused by BZ?<br>**Is there anything you do to avoid or reduce risks related to overdose when using BZs with opioids?**<br>➤ Trying one before buying<br>➤ Buying tablets in sealed packets<br>➤ Visually inspecting the thickness of the tablet<br>➤ Time between use<br>➤ Using with other people<br>➤ Carry naloxone<br>**What things would reduce the risk and/or help you reduce frequency of taking benzos and opioids?**<br>➤ Harm reduction advice – is any advice given currently? If so, what?<br>➤ Prescription of benzos from GP<br>➤ Overdose prevention centres<br>➤ Drug safety checking and information about what's in drugs |
| 6. | **Sleep Apnoea** | **How would you describe your sleep in general?**<br>➤ Do you wake up sometimes feeling as if you are choking (gasping for breath)? / Has anyone seen you stop breathing during your sleep? (explore /check if different from gouching out); if they use terms like shallow breathing or similar, ask them to describe in more detail (e.g. breathing frequency slows, or they feel like they are taking in less air)<br>➤ Does this happen more when you have taken a benzo?<br>➤ How does your drug use affect your sleep? (ask about daytime and night-time sleep). Probe for whether they experience daytime sleepiness / daytime sleep and drug use.<br>➤ Have you woken yourself up with snoring? Is this worse when you have taken a benzo? |
| 7. | **Close** | **Retrace topic guide to complete a mop up of any missed or unclear topics**<br>**That is the end of my questions. Before we finish is there anything I haven't covered today that you** |
|  |  | **would like to add?**<br>➤ Check in to see if participant is okay<br>➤ End of audio recording<br>➤ Thanks for participating<br>➤ Provide or explain how to access £10 as a thank you for their time. |
| **END OF INTERVIEW** |  |  |

## Footnotes

1 small round chocolate sweets covered in a colourful sugar shell – similar to M&Ms

2 For this study a near-fatal overdose was defined as receiving treatment from a paramedic, police officer, friend or family member (e.g. resuscitation, naloxone rescue) and/or hospital admission for overdose. It does not include events where people report prolonged periods of unconsciousness (*“a heavy gouch”*) without intervention.

